# Altered lower-limb kinematics and joint moments during high-impact tasks after hip and knee arthroplasties

**DOI:** 10.64898/2026.01.29.26345125

**Authors:** Bernard X.W. Liew, Fatemeh Farhadi, Leiming Gao, Stephen McDonnell, Wenxing Guo, Zainab Altai, Ahmed Soliman, Stefan Maas, Nelson Cortes

**Affiliations:** School of Sport, Rehabilitation and Exercise Sciences, University of Essex, Colchester, Essex, United Kingdom; University of Michigan and Shanghai Jiao Tong University Joint Institute, Shanghai Jiao Tong University, Shanghai, China; Department of Engineering, Nottingham Trent University; Division of Trauma and Orthopaedic Surgery, University of Cambridge, Addenbrooke’s Hospital; School of Mathematics, Statistics and Actuarial Science, University of Essex, Colchester, Essex, United Kingdom; Institute of Public Health and Wellbeing, University of Essex, Colchester, Essex, United Kingdom; Department of Engineering, Faculty of Science, Technology and Medicine, University of Luxembourg, Luxembourg; Department of Bioengineering, College of Engineering and Computing, George Mason University, VA USA

**Keywords:** Arthroplasty, Sports, Biomechanics, Running, Side-step cutting, Hopping, Return to sport, Total hip arthroplasty, Total knee arthroplasty

## Abstract

High-impact physical activity delivers musculoskeletal and systemic health benefits yet remains controversial for people with hip or knee arthroplasties due to concerns about implant loading and longevity. This study provides the first three-dimensional kinematic and kinetic characterisation of high-impact tasks in high-functioning adults with total hip arthroplasty (THA), total knee arthroplasty (TKA), or unicompartmental knee arthroplasty (UKA), compared with healthy controls. High-functioning adults with a joint arthroplasty (THA=11, TKA=4, UKA=3) participated. Healthy comparison data (n=70) were obtained from prior studies adopting the same analytical framework. They completed running, 45° change of direction (COD), countermovement jumps (CMJs), and unilateral/bilateral hopping. An 8-segment biomechanical model was used to quantify 3D joint angles and internal moments. These were time-normalised and modelled using Generalized Additive Models with covariates for group, age, and task intensity. THA had greater hip adduction angles during running 3.65° (95%CI 0.73°, 6.57°) and COD45 14.12° (95%CI 3.22°, 25.02°), compared to controls. THA participants recruited greater hip extensor moment during loading response in running, propulsion during CMJ, and in unilateral and bilateral hops, compared to controls. People with a TKA and UKA exhibited a greater internal rotation angle during running, without a concomitant increase in moments at initial contact and toe-off, compared to healthy controls. Also, TKA and UKA participants experienced reductions in knee extensor moments across all high-impact tasks, such as a 1.86 to 1.89 Nm/kg reduction during running. High-functioning individuals with a joint arthroplasty can perform demanding tasks, but often through movement strategies that may compromise long-term implant durability.

## Introduction

In the United Kingdom (UK) alone, 108,558 and 116,845 primary hip and knee arthroplasties, respectively, were undertaken in 2023 (Registry, 2024). The demand for joint arthroplasty is projected to increase by 40% by 2060 (Matharu et al., 2022), and patients receiving a joint arthroplasty are becoming younger (Kurtz et al., 2009). Younger patients are at greater risk of needing a revision of joint arthroplasty surgery (Bayliss et al., 2017), and are more active and have greater expectations of returning to vigorous physical activities (PA) compared to older adults (Ponzio et al., 2021).

People with a joint arthroplasty are traditionally advised to participate in low-impact PA (Lester et al., 2022; Sowers et al., 2023) and avoid vigorous, high-impact PA and sports, because of concerns regarding implant safety and longevity (Mooiweer et al., 2022). However, the World Health Organisation recommends that adults accumulate at least 75–150 min/week of vigorous-intensity aerobic PA (or an equivalent combination of moderate- and vigorous-intensity activity). Vigorous PA may involve high-impact tasks like running, jumping, landing, and change of direction tasks. High-impact PA have many superior health benefits over low-impact activities, such as being a better stimulus for bone adaptation (Allison et al., 2015). High-impact PA may be relevant to people with a joint arthroplasty, given that they are at an age, or growing into the age, where the risk of osteoporosis is greater (Clynes et al., 2020).

Observational studies have reported that high-impact activities do not increase the rates of joint arthroplasty revision (Kornuijt et al., 2022; Shah et al., 2025). However, these studies focus primarily on surgical revision as the outcome. Understanding the kinematics and kinetics of how people with a total hip or knee joint arthroplasty perform high-impact tasks is crucial for understanding the mechanical loads these patients place on their joints, and for informing potentially safer technique modification strategies. Many biomechanics studies in people with a joint arthroplasty have focused only on low-impact activities, such as walking (Dong et al., 2023; Lunn et al., 2019; Moyer et al., 2018). Far fewer studies have investigated the biomechanics of high-impact tasks in people with a joint arthroplasty (Harvey et al., 2024). Some studies investigated patients with an instrumented joint implant to quantify joint contact forces (Bergmann et al., 2016; Bergmann et al., 2014; Bergmann et al., 1993). These studies have not considered a wider range of high-impact activities beyond running, like hopping and countermovement jumps (CMJs) (Altai et al., 2024). Additionally, previous studies have not specifically included patients who have successfully returned to high-impact PA (Bergmann et al., 2016; Bergmann et al., 2014; Bergmann et al., 1993; Harvey et al., 2024).

This study aims to quantify the three-dimensional (3D) joint angles and moments during high-impact tasks in high-functioning people with hip or knee arthroplasties. We also aim to compare the 3D kinematics and kinetics of people with a joint arthroplasty against those of healthy people without a joint implant. The findings of the current study are intended to provide the biomechanical basis to inform future PA recommendations and to inform implant load-testing protocols.

## Methods

### Participants

Participants with a total hip arthroplasty (THA, n = 11), total knee arthroplasty (TKA, n = 4), or unicompartmental knee arthroplasty (UKA, n = 3) were enrolled. Participants were eligible if they were 1) between 18 years to 85 years old, 2) had undergone THA, TKA, or UKA surgery ≥ 1 year before enrolment, 3) self-reported returning to recreational or competitive high-impact PA, 4) reported no pain at the joint arthroplasty site, and 5) were free from any medical conditions that preclude safe participation in vigorous-intensity PA. All participants provided written informed consent, and the study was approved by the Health Research Authority Research Ethics Committee (IRAS 327418).

Data from healthy participants were obtained from prior studies (Altai et al., 2024; Liew et al., 2020). Forty-four (22 males, 22 females) healthy participants from Altai et al. (Altai et al., 2024) were included, with an average age, mass, and height of 38.8 (13.6) years, 68.2 (11.7) kg, and 1.7 (0.1) m, respectively. Twenty-six (16 males, 8 females) healthy participants from Liew et al. (Liew et al., 2020) were included, with an average age, mass and height of 22.0 (2.9) years, 68.4 (10.8) kg, and 1.8 (0.1) m, respectively.

### Experimental protocol

Participants with a joint arthroplasty performed all trials while wearing their usual comfortable running shoes and exercise attire. For running, participants performed at least three successful trials at a comfortable self-selected pace. For the COD45, participants ran at a comfortable pace before performing a 45° side-step cutting manoeuvre with their operated limb towards the contralateral direction. For running and COD45, a lead-up and tail-off distance of 10m was provided. For CMJs, participants performed three separate repetitions of maximal and submaximal (50% of perceived maximal) effort CMJs, each at the preferred descent depth, with the arms held in a “T” position (90° shoulder abduction). For hopping, participants performed two sets of bilateral and unilateral (operated-limb) hops at a fixed moderate intensity of 2.6 Hz using an auditory metronome. Participants were required to position their arms in a 90° abducted “T” position and hop continuously for 10 s. A minimum of 1 min rest was provided between each repetition of the task to avoid fatigue. Details of the experimental protocol for the data from the healthy participants can be found in the original publications (Altai et al., 2024; Liew et al., 2020), but also summarised in Table SM1 in the supplementary.

### Biomechanical model and processing

Retro-reflective markers were adhered to the trunk and lower extremities of each participant (see Supplementary Figure SM1). Marker trajectories were captured using 14 optical motion capture cameras (Vicon Ltd., 200 Hz), while ground reaction forces (GRF) were captured using two in-ground force plates (Kistler, 2000 Hz). Hip joint centres were calculated using a regression equation (Bell et al., 1989). Knee and ankle joint centres were calculated as the midpoint between the medial and lateral femoral condyles and malleoli, respectively (Pohl et al., 2010). An eight-segment trunk-lower extremity kinetic model was created in Visual 3D (Dempster, 1955; Hanavan, 1964).

Both marker trajectories and GRF were filtered at a common cut-off frequency of 18Hz, using a 4^th^ order, bidirectional, zero-lag Butterworth filter (Liew et al., 2016). Details of the biomechanical modelling protocol of the data from the healthy participants can be found in the original publications (Altai et al., 2024; Liew et al., 2020), and summarised in Table SM1 of the supplementary. For all studies, three-dimensional (3D) joint angles were calculated using a Cardan XYZ (flexion-extension, abduction-adduction, and axial rotation) rotation sequence (Cole et al., 1993). Internal joint moments were expressed within an orthogonal frame of reference of the proximal segment (Schache and Baker, 2007).

Joint angles and moments were segmented as follows: between consecutive initial contacts of the operated limb for hopping, between initial contact and toe-off for running and COD45 (i.e. stance), and between the onset of the model’s centre of mass (COM) descent and toe-off for CMJs. The time-varying joint angle and moment segments were time-normalised to 101 data points for all tasks. The intensity of the six tasks was quantified as mean COM speed (m/s) for running, mean COM approach speed (m/s) for COD45, stance duration (s) for hopping, and the jump height (m) for the CMJ, defined as the difference between peak COM height and standing COM height.

### Patient-reported outcome measures

The following data were collected as descriptive measures from participants with a joint arthroplasty. The Oxford Hip Score (OHS) and the Oxford Knee Score (OKS) were collected for participants with a hip or knee arthroplasty, respectively (score 0 (poorest function) to 48 (maximal function) (Dawson et al., 1998; Wylde et al., 2005)). The Hip Disability and Osteoarthritis Outcome Score (HOOS) and the Knee Disability and Osteoarthritis Outcome Score (KOOS) were collected for participants who underwent hip or knee arthroplasty, respectively (score 0 (extreme joint problems) to 100 (no joint problems) (Nilsdotter et al., 2003; Roos and Lohmander, 2003)). Lastly, the High-Activity Arthroplasty Score (HAAS) was collected (score 0 (poorest function) to 18 points (maximal function) (Talbot et al., 2010)).

### Statistical analysis

All statistical analyses were conducted using R software (version 4.4.2). There were a total of nine time-varying joint angles and nine time-varying joint moment dependent variables. The independent variables were groups (Healthy [reference], THA, TKA, UKA), age, task intensity, a group-by-cycle interaction effort, and a random subject intercept was included to account for the repeated measures of the study. The effects of the independent variables on the dependent variables were modelled using Generalized Additive Modelling (GAM) (Wood, 2017). The *mgcv (v1*.*9-3)* package was used for GAM to perform statistical inference, and *tidygam (v1*.*0*.*0)* was used for pairwise post-hoc contrast. A significant difference between groups was determined by a non-zero crossing of the 95% confidence interval (CI). In the main text, we focus on the report of hip variables for individuals with THA, and the knee variables for people with TKA or UKA.

## Results

Table 1 provides a summary of the descriptive characteristics of our participants with a joint arthroplasty. The supplementary material contains figures of the average predicted joint angle and moment waveforms, as well as the pairwise difference contrasts of the hip, knee, and ankle. Below, we report the predicted point estimate (95%CI) of the peak pairwise mean difference in the dependent variables between groups.

**Table 1.** Demographic characteristics of participants with a joint arthroplasty.

| Characteristic | THA N = 11<sup>l</sup> | TKA N = 4<sup>l</sup> | UKA N = 3<sup>l</sup> |
| --- | --- | --- | --- |
| Age (years) | 55.9 (9.8) | 63.7 (4.8) | 65.3 (2.5) |
| Sex |  |  |  |
| Female | 6 / 11 (55%) | 3 / 4 (75%) | 0 / 3 (0%) |
| Male | 5 / 11 (45%) | 1 / 4 (25%) | 3 / 3 (100%) |
| Height (m) | 1.7 (0.1) | 1.7 (0.1) | 1.8 (0.1) |
| Mass (kg) | 68.9 (10.7) | 70.9 (14.4) | 102.6 (25.7) |
| Tested side |  |  |  |
| Left | 5 / 11 (45%) | 3 / 4 (75%) | 1 / 3 (33%) |
| Right | 6 / 11 (55%) | 1 / 4 (25%) | 2 / 3 (67%) |
| Education |  |  |  |
| College or university | 4 / 11 (36%) | 1 / 4 (25%) | 0 / 3 (0%) |
| Higher or secondary or further education (A-levels, BTEC, etc.) | 1 / 11 (9.1%) | 0 / 4 (0%) | 0 / 3 (0%) |
| Post-graduate degree | 6 / 11 (55%) | 3 / 4 (75%) | 2 / 3 (67%) |
| Secondary school up to 16 years | 0 / 11 (0%) | 0 / 4 (0%) | 1 / 3 (33%) |
| Work |  |  |  |
| Full-time | 7 / 11 (64%) | 0 / 4 (0%) | 1 / 3 (33%) |
| Not working | 2 / 11 (18%) | 4 / 4 (100%) | 1 / 3 (33%) |
| Part-time | 2 / 11 (18%) | 0 / 4 (0%) | 1 / 3 (33%) |
| Oxford (0-48) | 47.2 (1.3) | 46.5 (2.4) | 45.0 (4.4) |
| K/HOOS Symptom (0-100) | 96.4 (5.5) | 72.3 (6.8) | 71.4 (0.0) |
| K/HOOS Pain (0-100) | 97.7 (4.7) | 95.1 (4.7) | 96.3 (6.4) |
| K/HOOS ADL (0-100) | 99.2 (1.5) | 98.5 (2.1) | 96.1 (6.8) |
| K/HOOS Sports (0-100) | 97.7 (4.2) | 88.8 (7.5) | 95.0 (8.7) |
| K/HOOS QOL (0-100) | 88.1 (12.6) | 90.6 (8.1) | 95.8 (7.2) |
| HAAS (0-18) | 17.1 (1.5) | 14.3 (1.0) | 13.7 (4.2) |
| Main Sport |  |  |  |
| Beach Volleyball | 1 / 11 (9.1%) | 0 / 4 (0%) | 0 / 3 (0%) |
| Dance | 0 / 11 (0%) | 1 / 4 (25%) | 0 / 3 (0%) |
| Dog Agility | 0 / 11 (0%) | 1 / 4 (25%) | 0 / 3 (0%) |
| Hockey/Pickle Ball/Hickes | 0 / 11 (0%) | 1 / 4 (25%) | 0 / 3 (0%) |
| Run | 10 / 11 (91%) | 1 / 4 (25%) | 1 / 3 (33%) |
| Tennis | 0 / 11 (0%) | 0 / 4 (0%) | 2 / 3 (67%) |
| Competitive level of main sport |  |  |  |
| Both recreational and competitive | 3 / 11 (27%) | 0 / 4 (0%) | 2 / 3 (67%) |
| Competitive | 1 / 11 (9.1%) | 1 / 4 (25%) | 0 / 3 (0%) |
| Recreational | 7 / 11 (64%) | 3 / 4 (75%) | 1 / 3 (33%) |
| Experience (years) in main sport |  |  |  |
| 1 to <3 | 1 / 11 (9.1%) | 0 / 4 (0%) | 0 / 3 (0%) |
| 10 or more | 9 / 11 (82%) | 4 / 4 (100%) | 2 / 3 (67%) |
| 5 to <10 | 1 / 11 (9.1%) | 0 / 4 (0%) | 1 / 3 (33%) |
| Weekly training frequency in main sport |  |  |  |
| 1-2 times per week | 5 / 11 (45%) | 2 / 4 (50%) | 1 / 3 (33%) |
| 3-4 times per week | 6 / 11 (55%) | 2 / 4 (50%) | 2 / 3 (67%) |

<sup>l</sup>Mean (SD); n / N (%)

### Joint angles

During running, people with a THA exhibited 9.31° (3.6°, 15.02°) greater hip flexion and 3.65° (0.73°, 6.57°) less hip abduction, and 5.75° (1.01°, 10.49°) greater hip external rotation at 23%, 42%, and 34% of the movement cycle, respectively, compared to controls (Table 2). People with TKA had 10.63° (5.13°, 16.12°) greater knee internal rotation at 0% compared to controls (Table 3). People with UKA had 30.77° (0.17°, 61.36°) less knee extension at 100% and 14.22° (8.14°, 20.30°) greater knee internal rotation at 100% compared to controls (Table 3). In CMJ, people with THA showed 17.32° (9.06°, 25.58°) greater hip flexion at 85% and 5.81° (1.92°, 9.69°) less hip internal rotation at 43% compared to controls (Table 2). People with TKA showed 30.87° (13.64°, 48.11°) greater knee flexion at 27% compared to controls (Table 3). During unilateral and bilateral hopping, hip kinematics did not differ between people with THA and control participants (Table 2). In unilateral hopping, people with TKA exhibited 6.08° (0.35°, 11.81°) less knee internal rotation at 31%, whilst people with UKA displayed 7.56° (0.48°, 14.64°) greater knee flexion at 59%, compared to controls (Table 3). In bilateral hopping, people with TKA had 9.24° (3.25°, 15.23°) greater knee flexion at 47%, and 6.34° (0.74°, 11.94°) greater knee external rotation at 23% compared to controls (Table 3). During COD45, people with THA had 14.12° (3.22°, 25.02°) less hip abduction at 61%, compared to controls (Table 2). No people with TKA knee angle differences were detected during COD45 (Table 3). People with UKA had 10.00° (1.62°, 18.38°) less knee abduction at 66%, compared to controls (Table 3).

**Table 2.** Point estimate (95% confidence interval) of the greatest magnitude of difference in joint angles for patients with a total hip arthroplasty.

| **Intensity** | **Joints** | **THA (°)** |
| --- | --- | --- |
| **Running** | Hip Flexion (+) | 9.31 (3.6, 15.02) @ 23% |
| **Running** | Hip Adduction (+) | 3.65 (0.73, 6.57) @ 42% |
| **Running** | Hip Internal (+) | -5.75 (-10.49, -1.01) @ 34% |
| **Running** | Hip Internal (+) | NS |
| **CMJ** | Hip Flexion (+) | 17.32 (9.06, 25.58) @ 85% |
| **CMJ** | Hip Adduction (+) | NS |
| **CMJ** | Hip Internal (+) | -5.81 (-9.69, -1.92) @ 43% |
| **Unilateral hopping** | Hip Flexion (+) | NS |
| **Unilateral hopping** | Hip Adduction (+) | NS |
| **Unilateral hopping** | Hip Internal (+) | NS |
| **Bilateral hopping** | Hip Flexion (+) | NS |
| **Bilateral hopping** | Hip Adduction (+) | NS |
| **Bilateral hopping** | Hip Internal (+) | NS |
| **COD45** | Hip Flexion (+) | NS |
| **COD45** | Hip Adduction (+) | 14.12 (3.22, 25.02) @ 61% |
| **COD45** | Hip Internal (+) | NS |
| **Abbreviations: CMJ – countermovement jump; COD45 – change of direction 45°, NS – no significance;** |  |  |

**Table 3.**
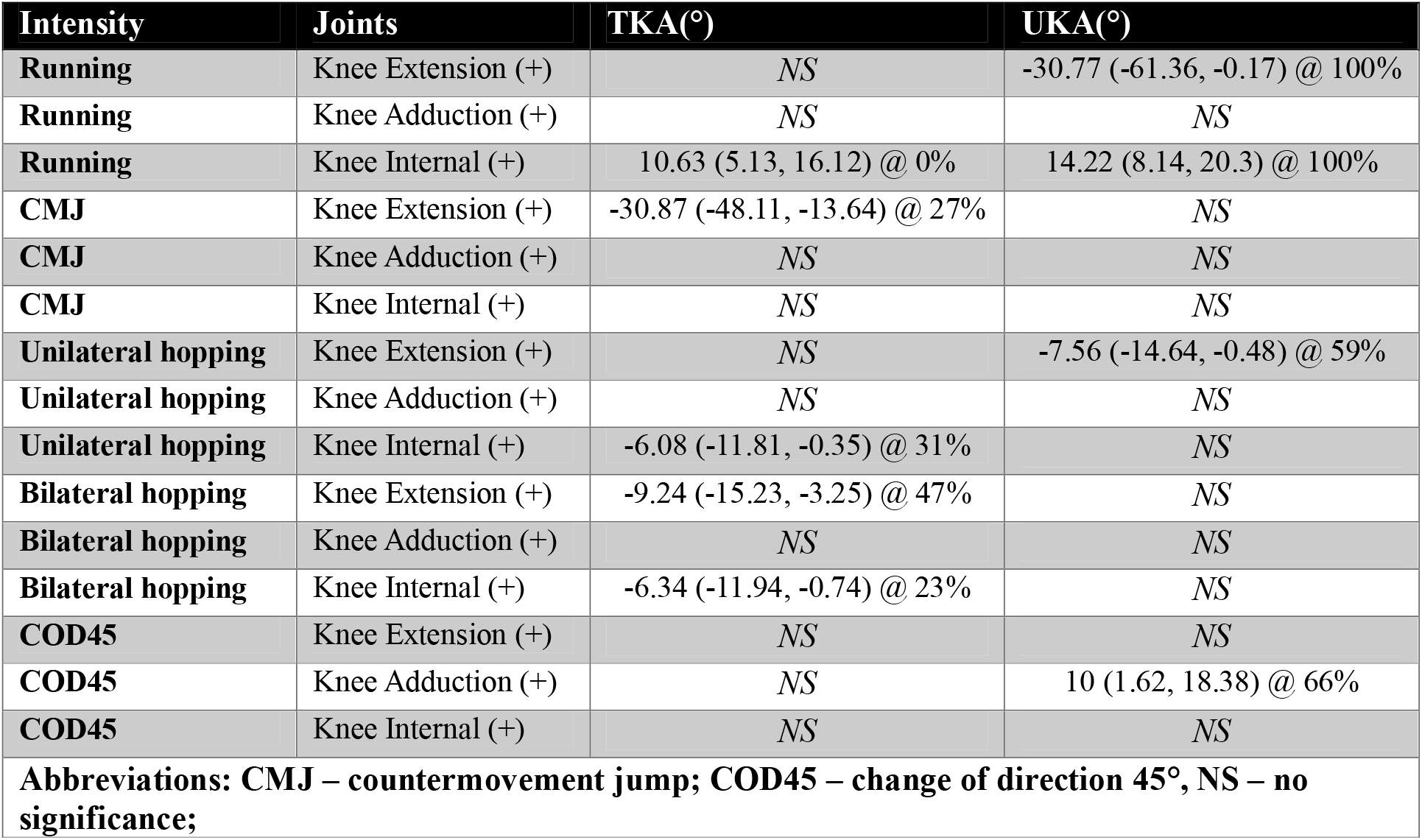
Point estimate (95% confidence interval) of the greatest magnitude of difference in joint angles for patients with a total or unicompartmental knee arthroplasty.

| Intensity | Joints | TKA(°) | UKA(°) |
| --- | --- | --- | --- |
| Running | Knee Extension (+) | NS | -30.77 (-61.36, -0.17) @ 100% |
| Running | Knee Adduction (+) | NS | NS |
| Running | Knee Internal (+) | 10.63 (5.13, 16.12) @ 0% | 14.22 (8.14, 20.3) @ 100% |
| CMJ | Knee Extension (+) | -30.87 (-48.11, -13.64) @ 27% | NS |
| CMJ | Knee Adduction (+) | NS | NS |
| CMJ | Knee Internal (+) | NS | NS |
| Unilateral hopping | Knee Extension (+) | NS | -7.56 (-14.64, -0.48) @ 59% |
| Unilateral hopping | Knee Adduction (+) | NS | NS |
| Unilateral hopping | Knee Internal (+) | -6.08 (-11.81, -0.35) @ 31% | NS |
| Bilateral hopping | Knee Extension (+) | -9.24 (-15.23, -3.25) @ 47% | NS |
| Bilateral hopping | Knee Adduction (+) | NS | NS |
| Bilateral hopping | Knee Internal (+) | -6.34 (-11.94, -0.74) @ 23% | NS |
| COD45 | Knee Extension (+) | NS | NS |
| COD45 | Knee Adduction (+) | NS | 10 (1.62, 18.38) @ 66% |
| COD45 | Knee Internal (+) | NS | NS |
| Abbreviations: CMJ – countermovement jump; COD45 – change of direction 45°, NS – no significance; |  |  |  |

### Joint moments

For running, people with a THA exhibited 0.74 (0.38, 1.10) Nm/kg less hip flexion moment at 65% of the movement cycle, 0.37 (0.12, 0.62) Nm/kg greater hip adduction moment at 76%, and 0.17 (0.04, 0.30) Nm/kg greater hip external rotation moment at 15%, compared to controls (Table 4). People with TKA or UKA exhibited significantly lower knee extension moment (1.86 to 1.89 Nm/kg) and lower knee abduction moment (0.67 to 0.68 Nm/kg) compared to controls (Table 5). For the CMJ, people with THA exhibited 0.58 (0.46, 0.69) Nm/kg greater hip extensor moment at 89%, 0.27 (0.20, 0.34) Nm/kg greater hip adduction moment at 82%, and 0.20 (0.14, 0.26) Nm/kg less hip external rotation moment at 73%, compared to controls (Table 4). People with a TKA and UKA had between 0.67 and 0.68 Nm/kg less knee extension moment, and between 0.10 and 0.14 Nm/kg less knee internal rotation moment, compared to controls (Table 5).

**Table 4.** Point estimate (95% confidence interval) of the greatest magnitude of difference in joint moments for patients with a total hip arthroplasty.

| **Intensity** | **Joints** | **THA (Nm/kg)** |
| --- | --- | --- |
| **Running** | Hip Flexion (+) | -0.74 (-1.1, -0.38) @ 65% |
| **Running** | Hip Adduction (+) | 0.37 (0.12, 0.62) @ 76% |
| **Running** | Hip Internal (+) | -0.17 (-0.3, -0.04) @ 15% |
| **Running** | Hip Internal (+) | -0.17 (-0.3, -0.04) @ 93% |
| **CMJ** | Hip Flexion (+) | -0.58 (-0.69, -0.46) @ 89% |
| **CMJ** | Hip Adduction (+) | 0.27 (0.2, 0.34) @ 82% |
| **CMJ** | Hip Internal (+) | 0.2 (0.14, 0.26) @ 73% |
| **Unilateral hopping** | Hip Flexion (+) | -0.67 (-0.81, -0.53) @ 32% |
| **Unilateral hopping** | Hip Adduction (+) | 0.51 (0.4, 0.63) @ 16% |
| **Unilateral hopping** | Hip Internal (+) | 0.16 (0.09, 0.23) @ 22% |
| **Bilateral hopping** | Hip Flexion (+) | -0.41 (-0.49, -0.34) @ 28% |
| **Bilateral hopping** | Hip Adduction (+) | 0.36 (0.3, 0.42) @ 21% |
| **Bilateral hopping** | Hip Internal (+) | 0.11 (0.09, 0.14) @ 23% |
| **COD45** | Hip Flexion (+) | 1.6 (0.9, 2.31) @ 8% |
| **COD45** | Hip Adduction (+) | NS |
| **COD45** | Hip Internal (+) | 0.63 (0.27, 0.99) @ 11% |
| **Abbreviations: CMJ – countermovement jump; COD45 – change of direction 45°, NS – no significance;** |  |  |

**Table 5.**
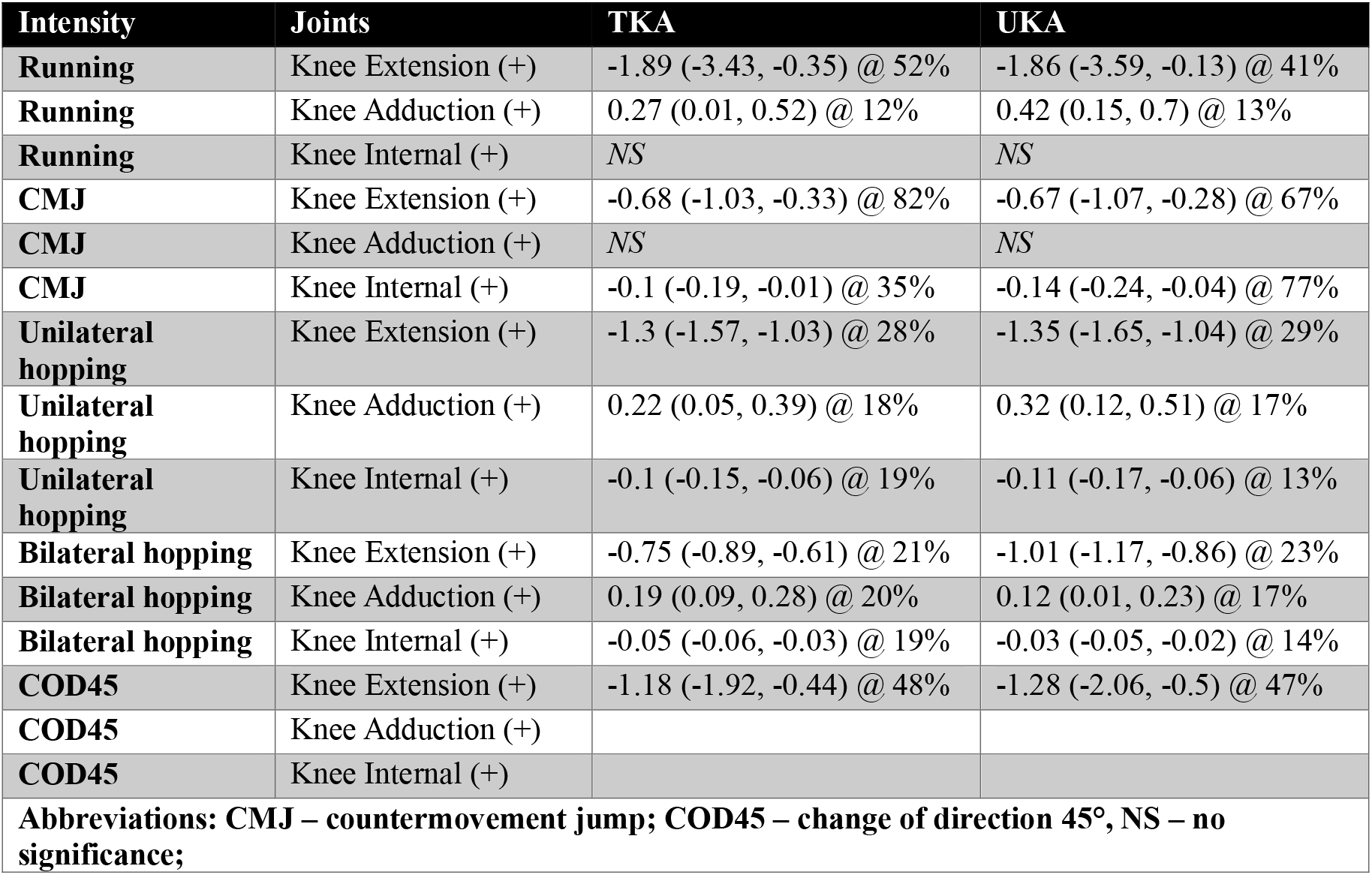
Point estimate (95% confidence interval) of the greatest magnitude of difference in joint moments for patients with a total or unicompartmental knee arthroplasty.

| **Intensity** | **Joints** | **TKA** | **UKA** |
| --- | --- | --- | --- |
| **Running** | Knee Extension (+) | -1.89 (-3.43, -0.35) @ 52% | -1.86 (-3.59, -0.13) @ 41% |
| **Running** | Knee Adduction (+) | 0.27 (0.01, 0.52) @ 12% | 0.42 (0.15, 0.7) @ 13% |
| **Running** | Knee Internal (+) | *NS* | *NS* |
| **CMJ** | Knee Extension (+) | -0.68 (-1.03, -0.33) @ 82% | -0.67 (-1.07, -0.28) @ 67% |
| **CMJ** | Knee Adduction (+) | *NS* | *NS* |
| **CMJ** | Knee Internal (+) | -0.1 (-0.19, -0.01) @ 35% | -0.14 (-0.24, -0.04) @ 77% |
| **Unilateral hopping** | Knee Extension (+) | -1.3 (-1.57, -1.03) @ 28% | -1.35 (-1.65, -1.04) @ 29% |
| **Unilateral hopping** | Knee Adduction (+) | 0.22 (0.05, 0.39) @ 18% | 0.32 (0.12, 0.51) @ 17% |
| **Unilateral hopping** | Knee Internal (+) | -0.1 (-0.15, -0.06) @ 19% | -0.11 (-0.17, -0.06) @ 13% |
| **Bilateral hopping** | Knee Extension (+) | -0.75 (-0.89, -0.61) @ 21% | -1.01 (-1.17, -0.86) @ 23% |
| **Bilateral hopping** | Knee Adduction (+) | 0.19 (0.09, 0.28) @ 20% | 0.12 (0.01, 0.23) @ 17% |
| **Bilateral hopping** | Knee Internal (+) | -0.05 (-0.06, -0.03) @ 19% | -0.03 (-0.05, -0.02) @ 14% |
| **COD45** | Knee Extension (+) | -1.18 (-1.92, -0.44) @ 48% | -1.28 (-2.06, -0.5) @ 47% |
| **COD45** | Knee Adduction (+) |  |  |
| **COD45** | Knee Internal (+) |  |  |
| **Abbreviations: CMJ – countermovement jump; COD45 – change of direction 45°, NS – no significance;** |  |  |  |

For unilateral hopping, people with THA experienced 0.67 (0.53, 0.81) Nm/kg greater hip extension moment at 32% of the movement cycle, 0.51 (0.40, 0.63) Nm/kg less hip abduction moment at 16%, and 0.16 (0.09, 0.23) Nm/kg less hip external rotation moment at 22%, compared to controls (Table 4). People with TKA and UKA both experienced 1.30 to 1.35 Nm/kg lower knee extension moment, 0.22 to 0.32 Nm/kg less knee abduction moment, and 0.10 to 0.11 Nm/kg less knee internal rotation moment, compared to controls (Table 5).

For bilateral hopping, people with THA experienced 0.41 (0.34, 0.49) Nm/kg less hip flexion moment at 28%, 0.36 (0.30, 0.42) Nm/kg less hip abduction moment at 21%, and 0.11 (0.09, 0.14) Nm/kg greater hip internal rotation moment at 23%, compared to controls (Table 4). People with TKA experienced lower knee extension moment (0.75 to 1.01 Nm/kg), greater knee abduction moment (0.12 to 0.19 Nm/kg), and 0.03 to 0.05 Nm/kg greater knee external rotation moment compared to controls (Table 5).

For COD45, people with THA experienced 1.60 (0.90, 2.31) Nm/kg less hip extension moment at 8% and 0.63 (0.27, 0.99) Nm/kg less hip external rotation moment at 11%, compared to controls (Table 4). People with TKA or UKA both experienced lower knee extension moment (1.18–1.28 Nm/kg) compared to controls (Table 5).

## Discussion

There is now a greater awareness that a return to high-impact sports may be possible after a joint arthroplasty. Most research on high-impact sports after joint arthroplasty relies on self-reported data (Kornuijt et al., 2022; Shah et al., 2025). To our knowledge, this is the first study to quantify the 3D joint angles and moments of the lower limbs in high-functioning people with hip or knee implants. Our findings collectively suggest that people with a joint arthroplasty who have successfully returned to high-impact PA may adopt biomechanical strategies that could compromise implant longevity.

Gait alterations have a significant influence on joint contact force magnitude and the direction of force application to the joints (Foucher et al., 2009; Wesseling et al., 2016). Alterations in joint force magnitudes and direction can negatively influence implant loosening and accelerate implant wear (Foucher et al., 2009). Individuals with a THA had greater hip adduction during running and COD45, compared to controls. Previous studies reported greater hip adduction angles during walking, which can be attributed to a reduced hip abductor strength (Fujita et al., 2025; Lunn et al., 2019). A greater hip adduction angle may increase the hip joint contact force magnitude (Wesseling et al., 2016) and the risk of edge loading during activities requiring greater hip flexion (van Arkel et al., 2013). This could increase contact stress and accelerate implant wear (Mellon et al., 2011; Underwood et al., 2012). Given that our participants with a THA performed running and CMJ with greater hip flexion and hip adduction than controls, this movement strategy may negatively increase implant wear.

People with THA exhibited greater hip extensor moment during shock absorption in running, the propulsive phase of CMJ, and during unilateral and bilateral hops, compared to controls. This is consistent with previous studies on walking, which reported that hip extensor moment or power increased from pre-to post-operation after a THA (Queen et al., 2019; Zhong et al., 2025). Another study reported greater peak hip power generation and absorption in high-functioning patients with THA compared to controls (Lunn et al., 2019). Our findings are unlikely to be attributed to the typical distal-to-proximal (i.e. less ankle, greater hip) shift in muscle function during age associated with ageing (Kulmala et al., 2014; Liew et al., 2025). This is because we included age as a covariate during statistical inference. Notably, participants with a THA performed running, unilateral and bilateral hopping with greater ankle extensor moment, compared to controls. However, a greater hip extensor moment could be in compensation for a reduction in knee extensor moment in people with THA (Wada et al., 2025). This suggests that rehabilitation of the quadriceps group may be required to reduce the reliance of the hip extensors, which may otherwise increase hip implant load and wear.

Both people with TKA and UKA ran, jumped, and hopped with reduced knee extension moments. This suggests knee extensor offloading appeared to be a common pattern observed in people with TKA and UKA during high-impact sports. In turn, people with TKA and UKA appeared to rely more on their hip extensors. One study reported that people with a TKA ran with a lower peak knee extensor moment, compared to controls (Harvey et al., 2024). Previous studies have reported that peak knee flexion angle and knee extensor moment were lower in people with TKA compared to controls during downslope treadmill walking (Wen et al., 2022). Even though downslope walking does not represent high-impact activities, it still requires a greater knee extensor moment than level-ground walking (Lay et al., 2006), which may partially mirror the mechanical demand of high-impact activities. The role of the knee is particularly important for shock absorption during high-impact activities, such as running, which requires a greater knee eccentric extensor moment (Schache et al., 2015).

People with a TKA and UKA exhibited a greater internal knee rotation angle during running, without a concomitant increase in knee internal moments, compared to healthy controls. Previous joint simulator studies have reported that a greater axial knee range of motion (ROM) with loading resulted in greater knee implant wear (Johnson et al., 2001; McEwen et al., 2005).

However, another study reported that the biggest gait biomechanics contributor to knee implant wear was due to increases in the peak magnitude of the knee sagittal plane ROM (Mell et al., 2020). In the present study, the CMJ and bilateral hopping resulted in sustained increases in knee flexion angle in people with a TKA, compared to controls. CMJ and hopping are increasingly being recognised as safe and effective exercises to be prescribed for older adults to improve neuromuscular function and bone density (Allison et al., 2015; Vetrovsky et al., 2019). The short- and long-term safety of such high-impact exercises in people with a knee joint implant is unclear. Nevertheless, the present findings suggest that such exercises should be accompanied by supervision to ensure optimal exercise technique.

The present study has several limitations. First, our sample size of people with a joint arthroplasty was small. The proportion of people with a joint arthroplasty who have returned to high-impact activities after surgery is likely to be still low, given that predominant post-operative clinical advice still recommends avoiding high-impact activities. Second, we did not collect data on age-matched healthy controls. This was because this project was conducted as part of a funded project focused on understanding how different physical activities affect the health of joint implants. However, we were able to use existing datasets on healthy participants for comparison, whilst statistically controlling for age and performance confounders. Third, we included a heterogeneous sample of participants with different implant manufacturers, which impacts the implant’s design, geometry, materials, and surgical approach. Future studies are warranted to understand how different implant and surgical factors may alter the biomechanics of high-impact sports, to better align surgical decision-making with participants’ lifestyle demands.

## Conclusions

Biomechanical studies are an essential companion to epidemiological studies for evaluating the safety of high-impact activities after a joint arthroplasty. High-functioning adults with hip or knee arthroplasty can perform high-impact tasks, but do so in a way that may negatively influence the longevity of the implants. The present study provides joint kinematics and kinetics data that may serve as input to enhanced implant stress testing protocols that reflect a physically demanding lifestyle after a joint arthroplasty. In addition, this study provides a foundation to develop novel physical rehabilitation approaches that provide patients with the benefits of high-impact activities without deleterious effects associated with participation.

## Data Availability

All data produced in the present study are available upon reasonable request to the authors

## Ethics approval and consent to participate

All participants provided written informed consent, and the study received approval from the Health Research Authority Research Ethics Committee (IRAS 327418).

## Acknowledgements

We thank Ms Mahbi Razavi for helping with data collection

## Authors’ contributions

**Conceptualisation:** Bernard Liew, Leiming Gao, Stephen McDonnell, Wenxing Guo, Ahmed Soliman, Stefan Maas, Nelson Cortes**; Funding Acquisition:** Bernard Liew, Leiming Gao, Stephen McDonnell, Wenxing Guo, Ahmed Soliman, Stefan Maas, Nelson Cortes**; Data collection:** Bernard Liew, Fatemeh Farhadi; **Data processing:** Bernard Liew, Fatemeh Farhadi, Zainab Altai; **Statistical modelling:** Bernard Liew, Wenxing Guo; **Validation**: All authors**; Visualisation**: Bernard Liew, Ahmed Soliman, Zainab Altai, Wenxing Guo; **Manuscript writing:** All authors; **Manuscript revision:** All authors; **Supervision:** Stefan Maas, Nelson Cortes

## Acknowledgements

Not applicable.

