## Supplementary material for "Altered lower-limb kinematics and joint moments during high-impact tasks after hip and knee arthroplasties"

**Journal category:** Original article

*Corresponding author

### Methodology summary

Table SM 1. Brief methodologies of included studies for healthy adults

|  | Altai et al. (Altai et al., 2024) | Liew et al. (Liew et al., 2020) |
| --- | --- | --- |
| Country | England | Australia |
| Inclusion criteria | 40 healthy adults, free from injuries or pain | 17 healthy adults, free from injuries or pain |
| Camera system | 16 cameras (Vicon, UK) | 18 cameras (Vicon T-series, Oxford Metrics, UK) |
| Experimental protocol | All – 3 successful trials each  Run – natural (typical jogging pace)  CMJ – maximal and medium effort, arms abducted “T” pose, self-selected depth  Hopping – vertical bilateral and unilateral (dominant, side used to kick ball), at 2.6Hz for 10s | 3 successful trials  COD45 – approach speed of 4 m/s, side-step cut with left leg to the right. |
| Camera sampling frequency | 200 Hz | 250Hz |
| Force platform system | 2 force plates (Kistler, Switzerland) | 4 force plates (AMTI, Watertown, MA, USA) |
| Force sampling frequency | 2000Hz | 1000Hz |
| Filtering | Markers – 18 Hz (2^nd^ Order, Butterworth, Zero lag)  GRF – 50 Hz (2^nd^ Order, Butterworth, Zero lag) | Markers – 12 Hz (4^th^ Order, Butterworth, Zero lag)  GRF – 12 Hz (4^th^ Order, Butterworth, Zero lag) |
| Footwear | Shod | |
| Surface | Overground | |
| Biomechanical model | Hip joint centre – Bell’s regression equation (Bell et al., 1989)  Knee joint centres – midpoint of medial and lateral femoral epicondyles  Ankle joint centre – midpoint of medial and lateral malleoli  Six degrees of freedom joints  Segment inertial and geometric properties were based on Dempster et al (1955)(Dempster, 1955) and Hanavan et al (1964) (Hanavan, 1964) | |

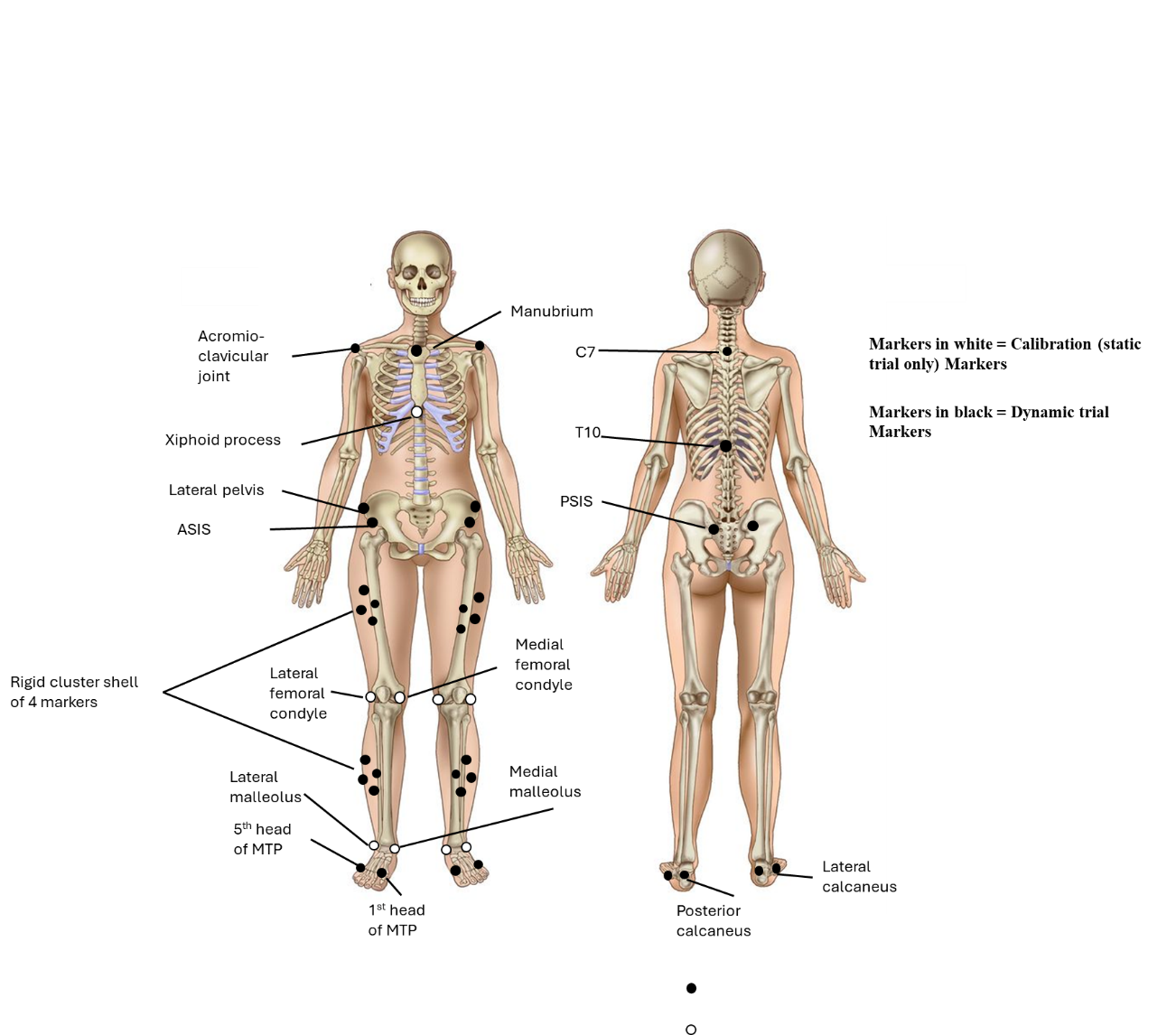

### Figure SM1. Marker placement protocol

### Joint angle predicted plots

Figures below reflect the time-varying waveforms for the various activities. Lines reflect the group average, whilst error clouds reflect 95% confidence intervals.

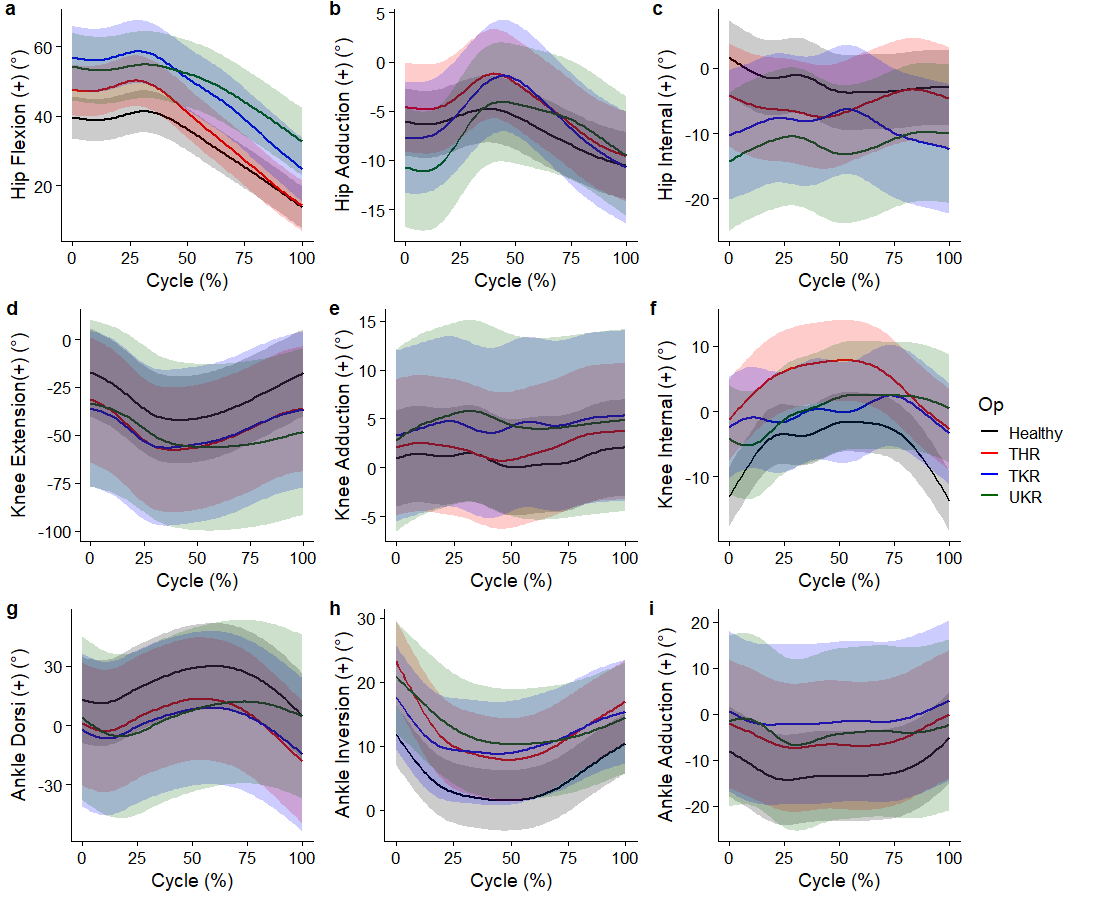

#### Figure SM2. Running – cycle reflects the stance phase

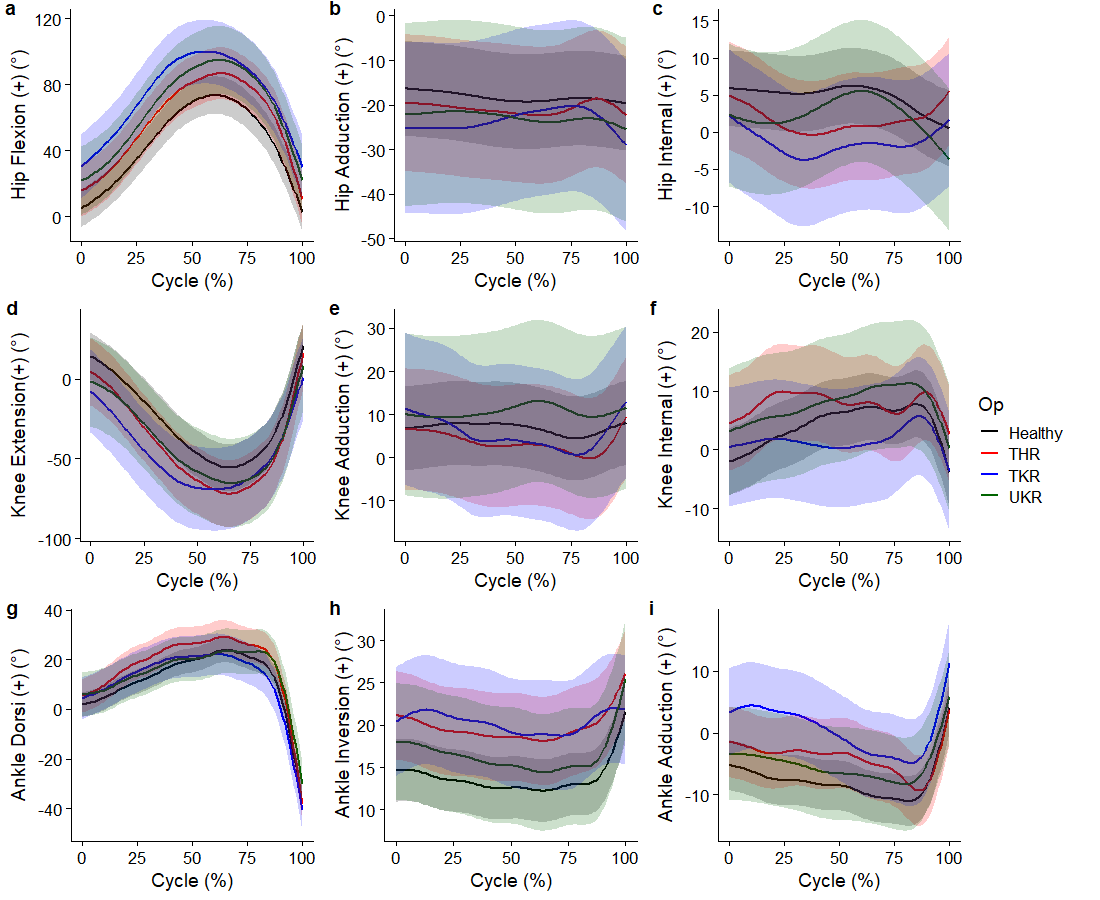

#### Figure SM3. Countermovement jumps – cycle reflects from the lowering of the centre of mass to toe-off

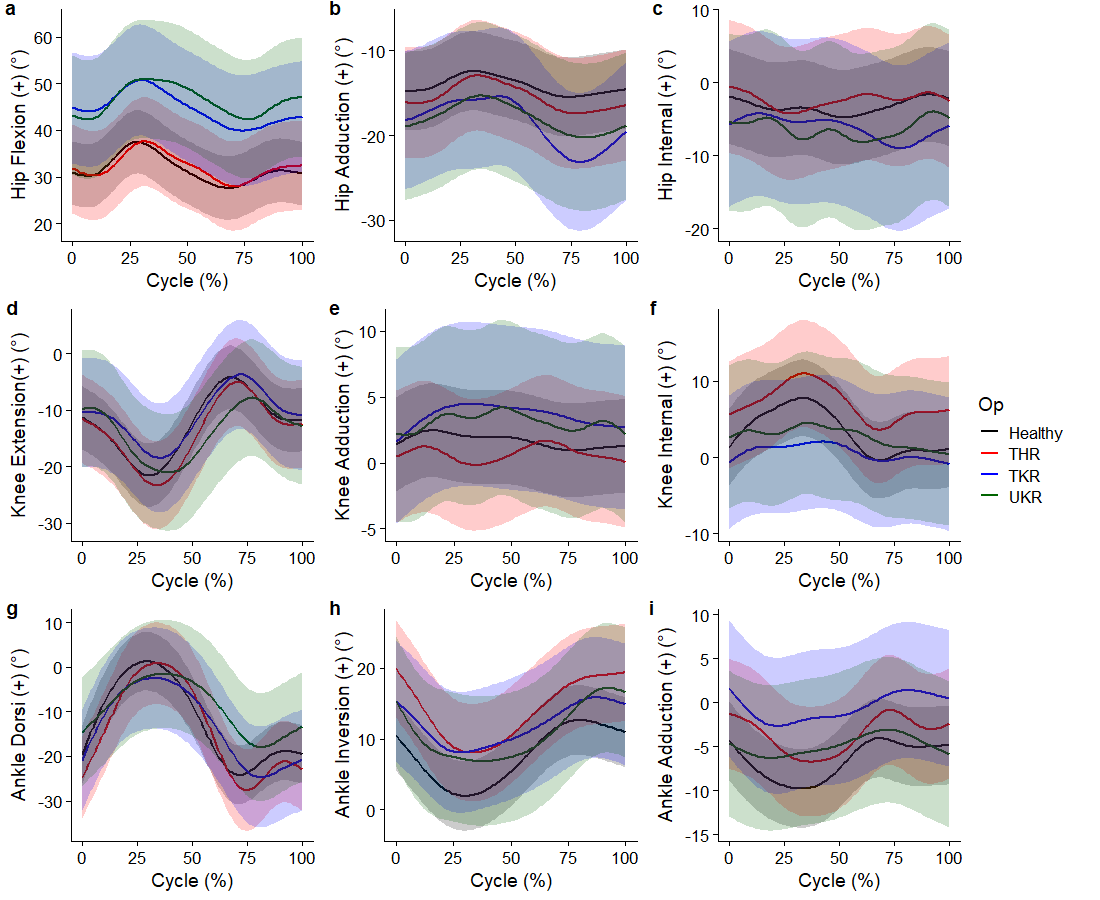

#### Figure SM4. Unilateral hopping – cycle reflects the stride phase

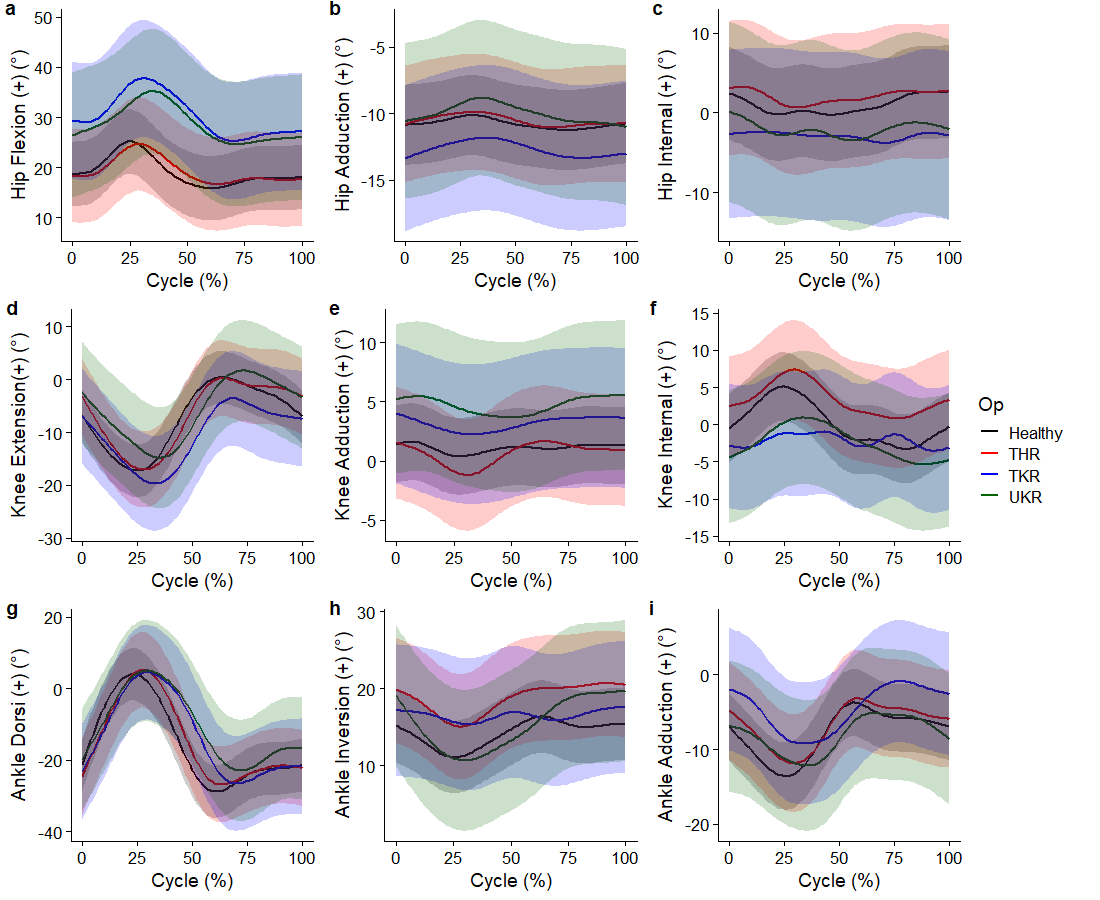

#### Figure SM5. Bilateral hopping – cycle reflects the stride phase

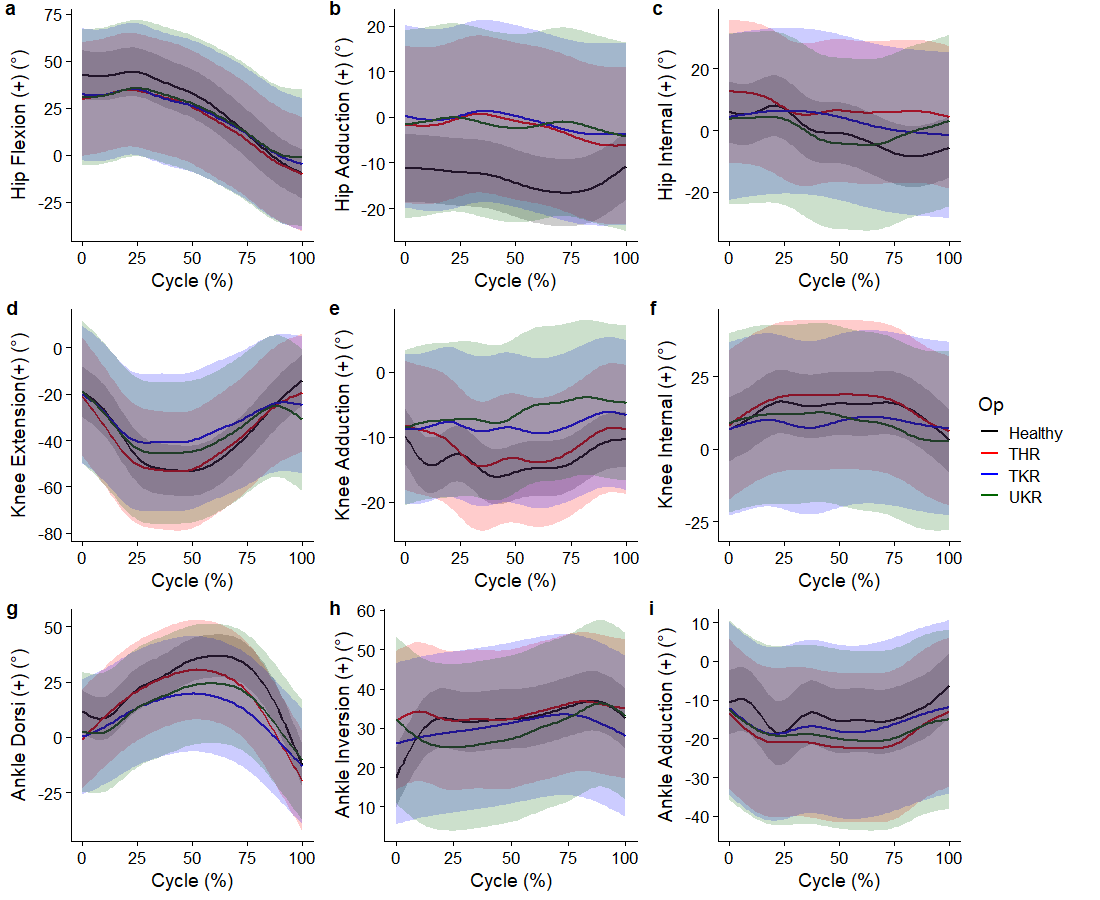

#### Figure SM6. Change of direction 45° – cycle reflects the stance phase

### Joint angle difference plots

Figures below reflect the time-varying waveforms of the pairwise difference between groups for each point on the movement cycle during various activities. Lines reflect the point estimate difference, whilst error clouds reflect 95% confidence intervals. Horizontal lines above waveforms reflect periods of statistically significant difference (i.e. non-zero cross of 95% confidence intervals).

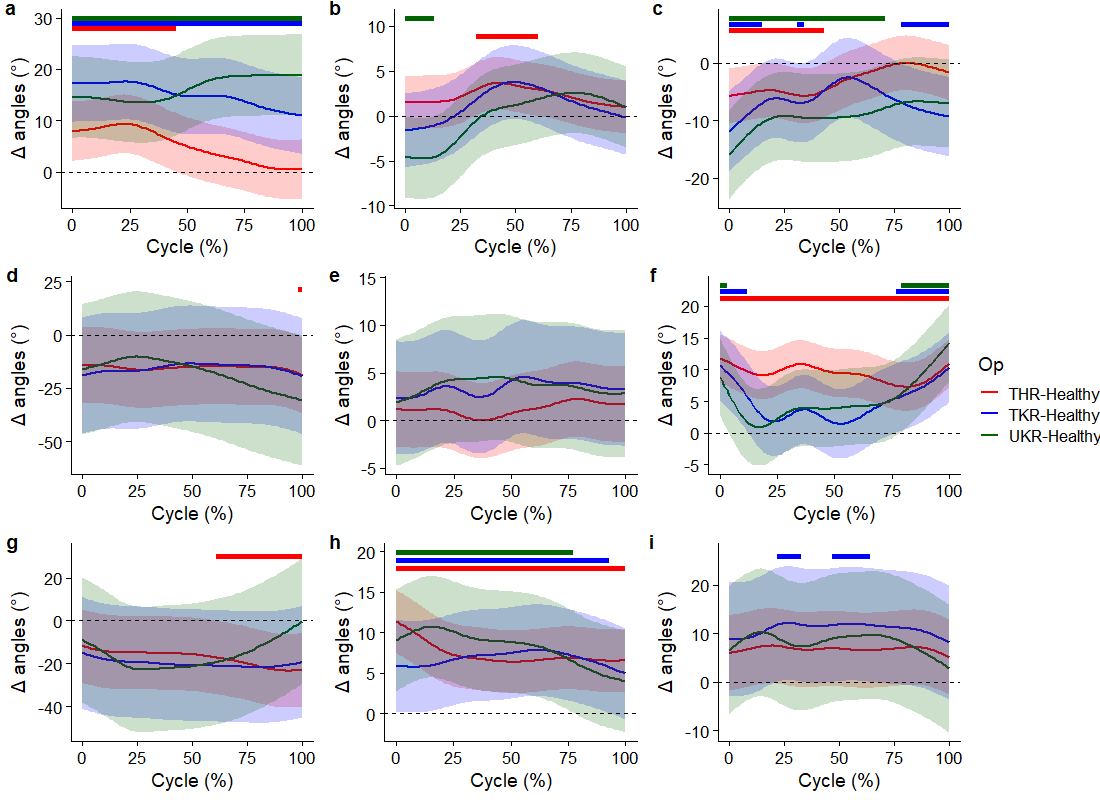

#### Figure SM7. Running – cycle reflects the stance phase.

(a) Hip flexion (+ = greater flexion in target), (b) Hip adduction (+ = greater adduction in target), (c) Hip internal rotation (+ = greater internal rotation in target), (d) Knee flexion (+ = greater extension in target), (e) Knee adduction (+ = greater adduction in target), (f) Knee internal rotation (+ = greater internal rotation in target), (g) Ankle Dorsi (+ = greater dorsiflexion in target), (h) Ankle inversion (+ = greater inversion in target), (i) Ankle Adduction (+ = greater adduction in target).

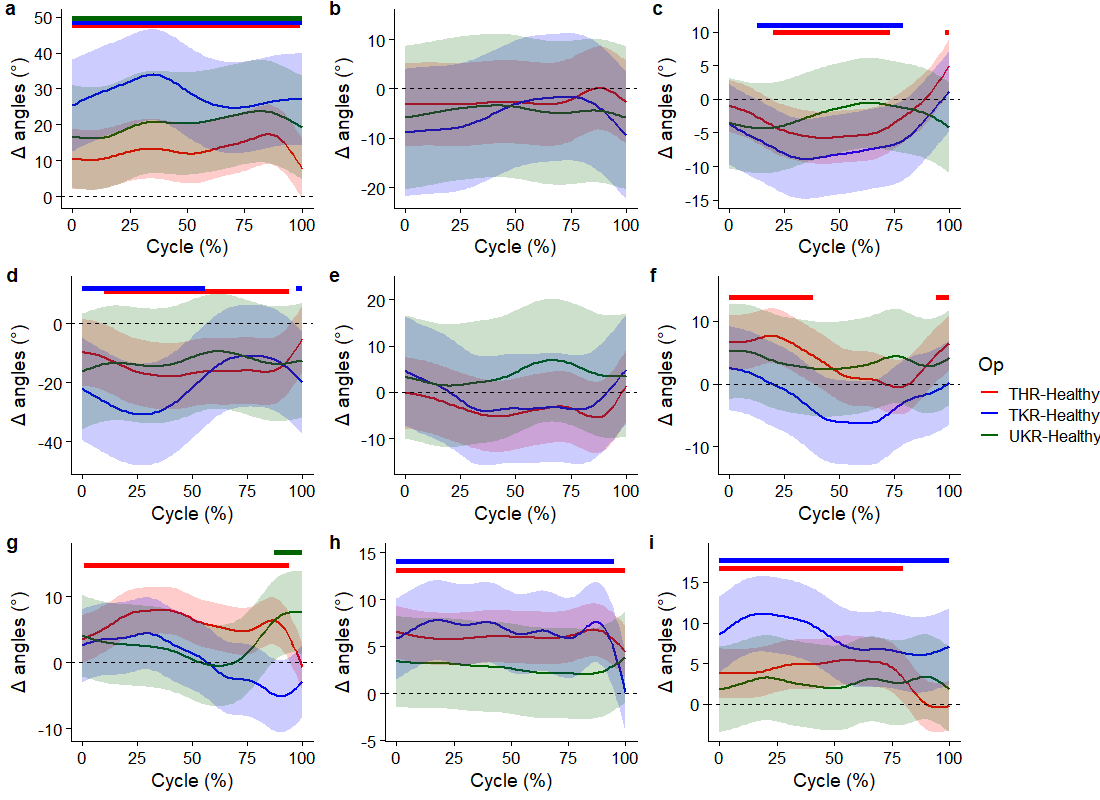

#### Figure SM8. Countermovement jumps – cycle reflects from the lowering of the centre of mass to toe-off

(a) Hip flexion (+ = greater flexion in target), (b) Hip adduction (+ = greater adduction in target), (c) Hip internal rotation (+ = greater internal rotation in target), (d) Knee flexion (+ = greater extension in target), (e) Knee adduction (+ = greater adduction in target), (f) Knee internal rotation (+ = greater internal rotation in target), (g) Ankle Dorsi (+ = greater dorsiflexion in target), (h) Ankle inversion (+ = greater inversion in target), (i) Ankle Adduction (+ = greater adduction in target).

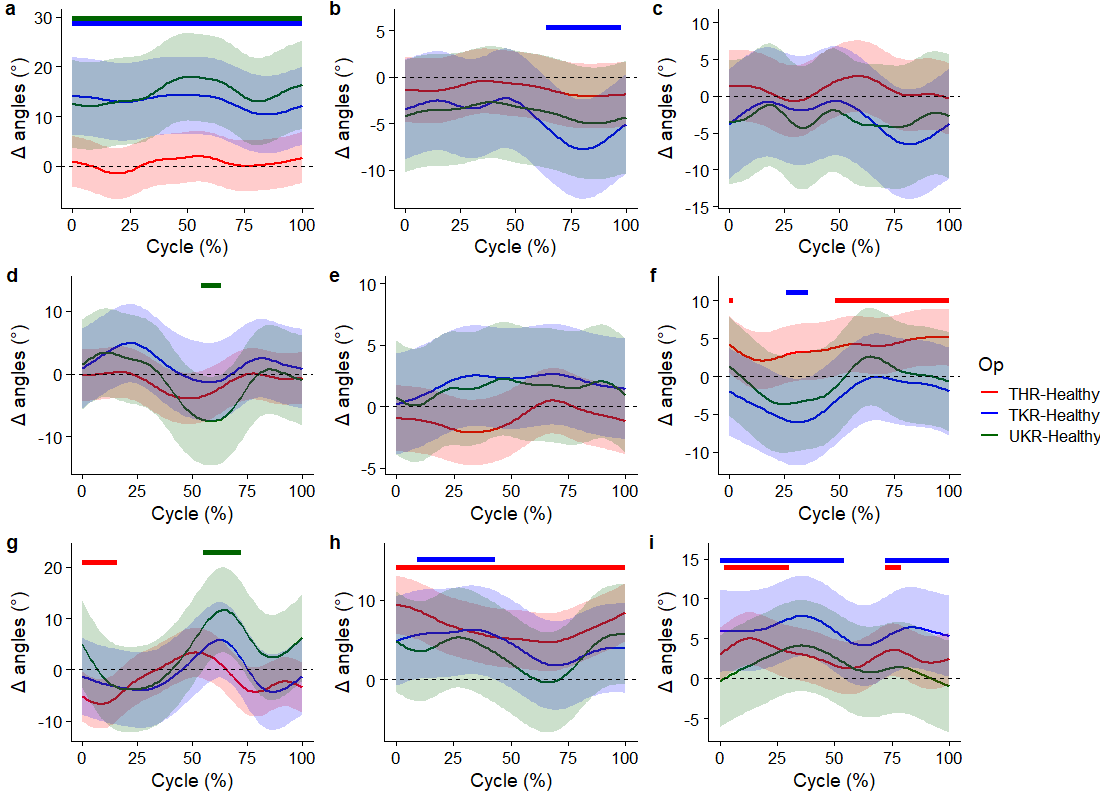

#### Figure SM9. Unilateral hopping – cycle reflects the stride phase

(a) Hip flexion (+ = greater flexion in target), (b) Hip adduction (+ = greater adduction in target), (c) Hip internal rotation (+ = greater internal rotation in target), (d) Knee flexion (+ = greater extension in target), (e) Knee adduction (+ = greater adduction in target), (f) Knee internal rotation (+ = greater internal rotation in target), (g) Ankle Dorsi (+ = greater dorsiflexion in target), (h) Ankle inversion (+ = greater inversion in target), (i) Ankle Adduction (+ = greater adduction in target).

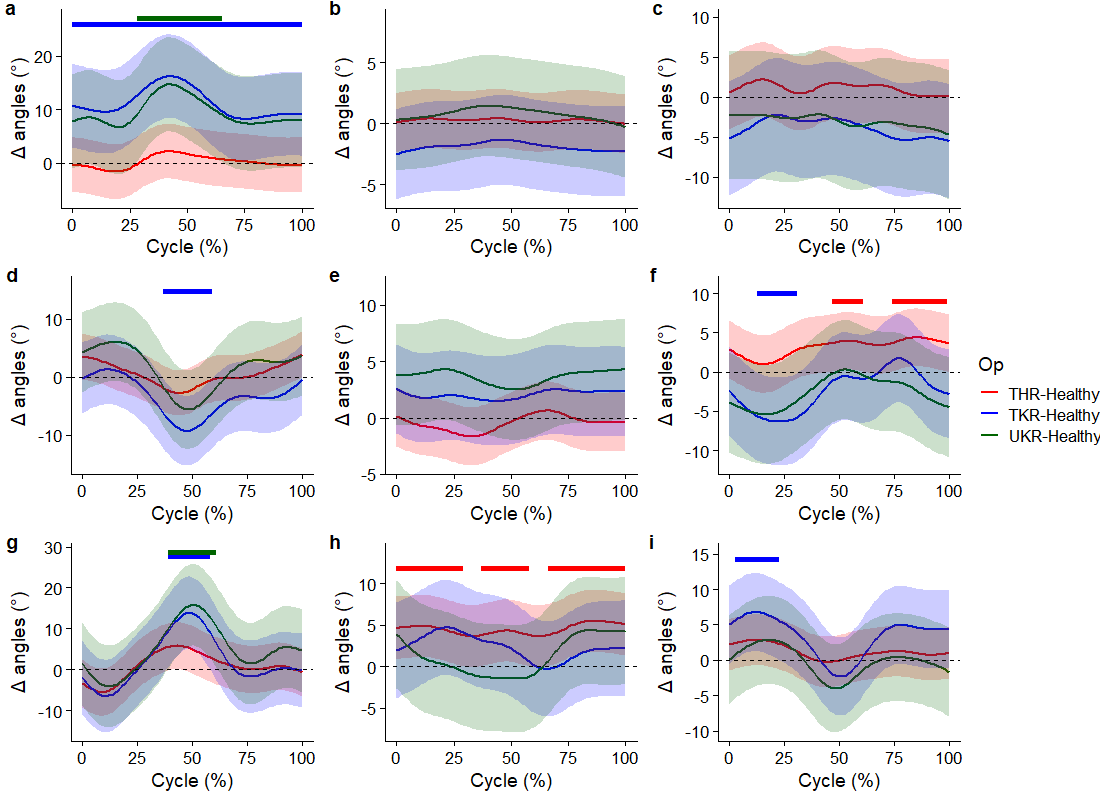

#### Figure SM10. Bilateral hopping – cycle reflects the stride phase

(a) Hip flexion (+ = greater flexion in target), (b) Hip adduction (+ = greater adduction in target), (c) Hip internal rotation (+ = greater internal rotation in target), (d) Knee flexion (+ = greater extension in target), (e) Knee adduction (+ = greater adduction in target), (f) Knee internal rotation (+ = greater internal rotation in target), (g) Ankle Dorsi (+ = greater dorsiflexion in target), (h) Ankle inversion (+ = greater inversion in target), (i) Ankle Adduction (+ = greater adduction in target).

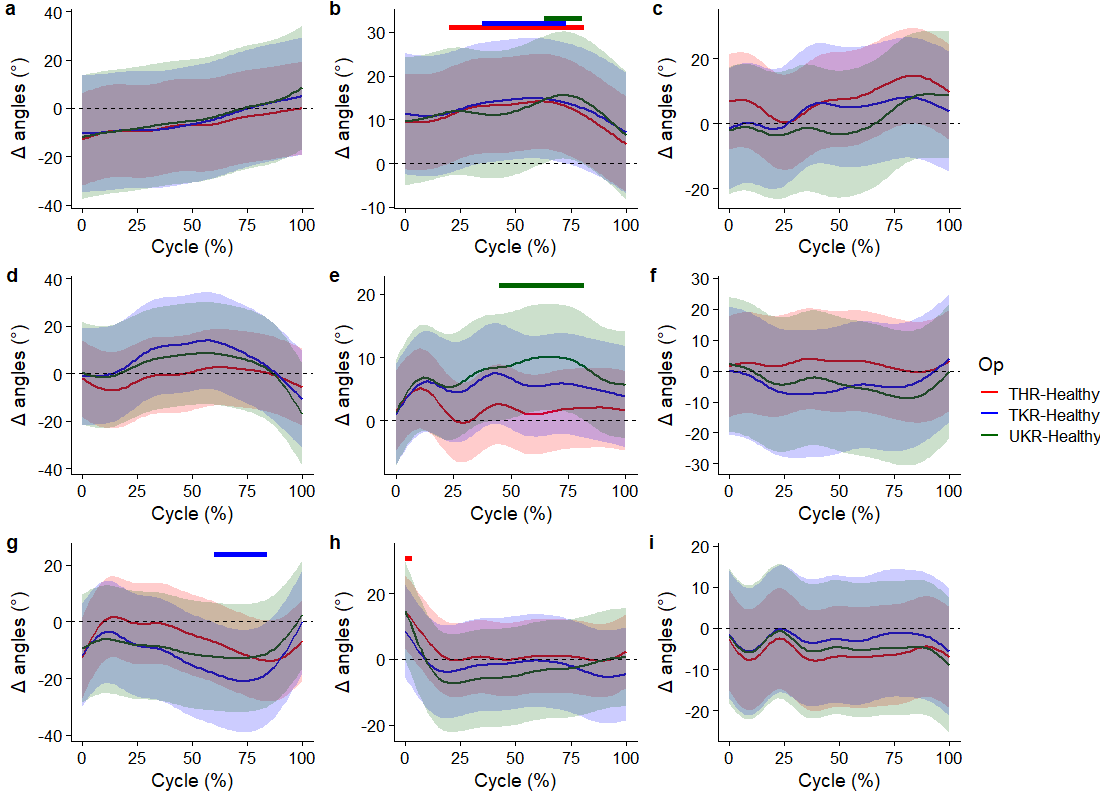

#### Figure SM11. Change of direction 45° – cycle reflects the stance phase

(a) Hip flexion (+ = greater flexion in target), (b) Hip adduction (+ = greater adduction in target), (c) Hip internal rotation (+ = greater internal rotation in target), (d) Knee flexion (+ = greater extension in target), (e) Knee adduction (+ = greater adduction in target), (f) Knee internal rotation (+ = greater internal rotation in target), (g) Ankle Dorsi (+ = greater dorsiflexion in target), (h) Ankle inversion (+ = greater inversion in target), (i) Ankle Adduction (+ = greater adduction in target).

### Joint moment predicted plots

Figures below reflect the time-varying waveforms for the various activities. Lines reflect the group average, whilst error clouds reflect 95% confidence intervals.

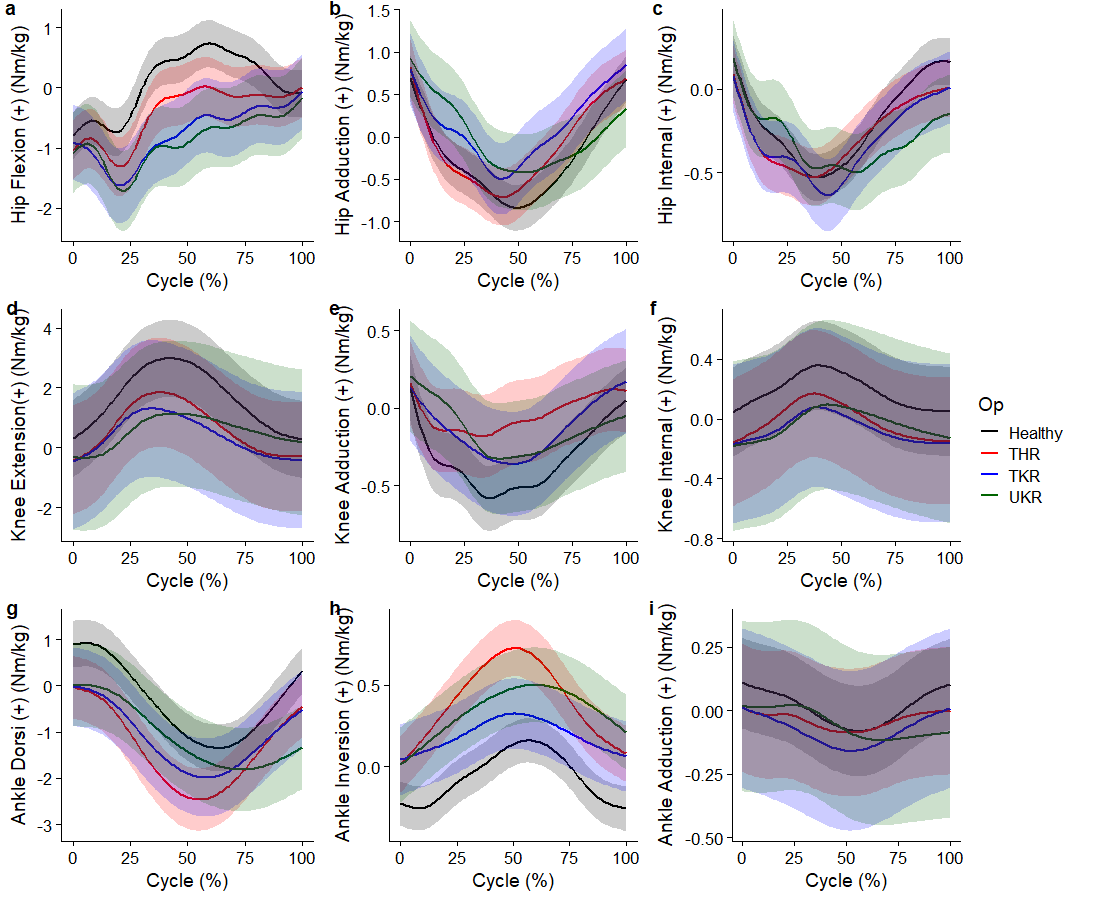

#### Figure SM12. Running – cycle reflects the stance phase

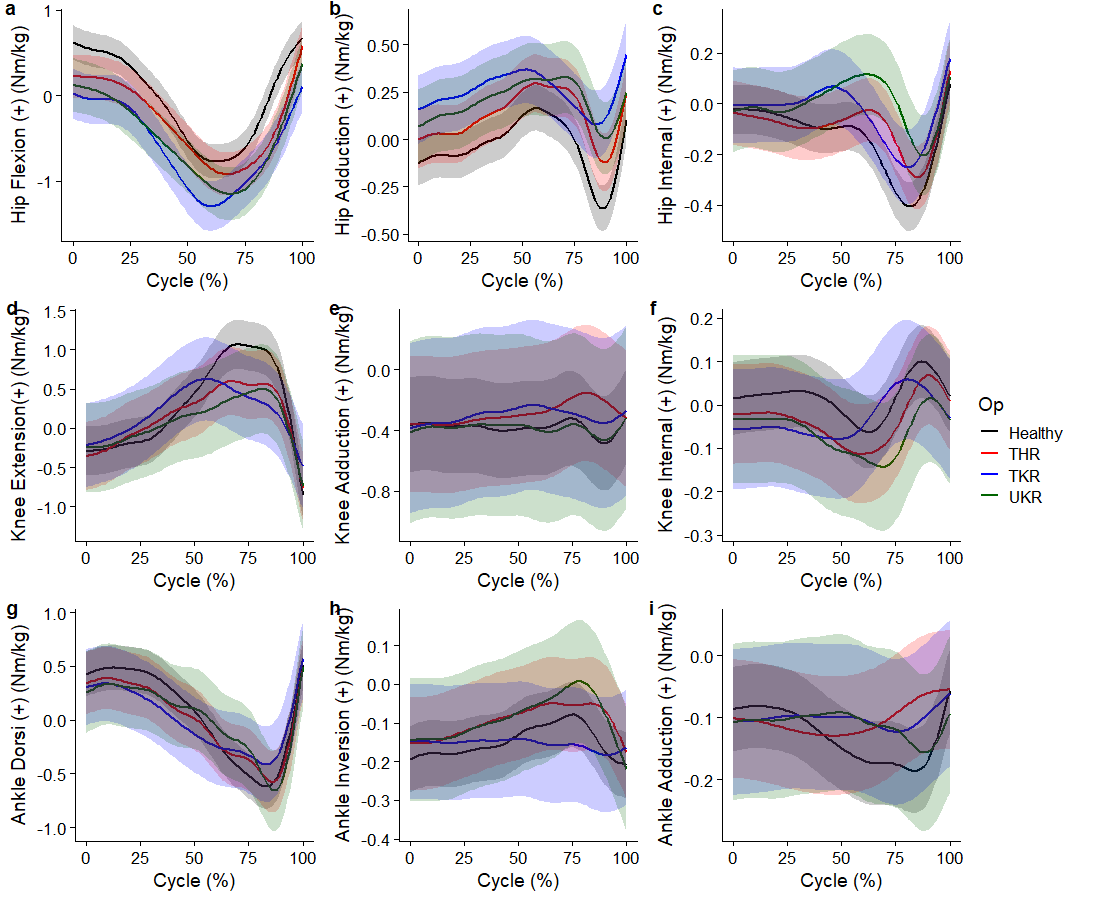

#### Figure SM13. Countermovement jumps – cycle reflects from the lowering of the centre of mass to toe-off

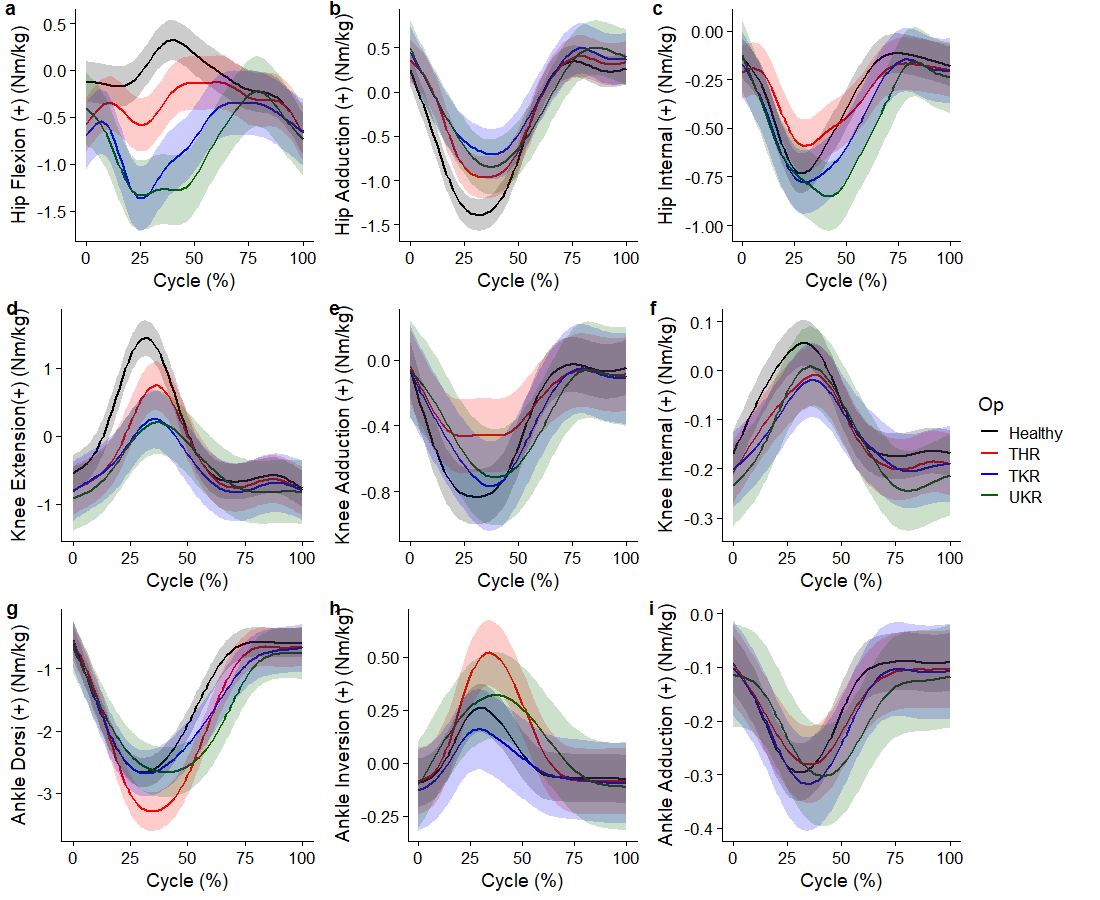

#### Figure SM14. Unilateral hopping – cycle reflects the stride phase

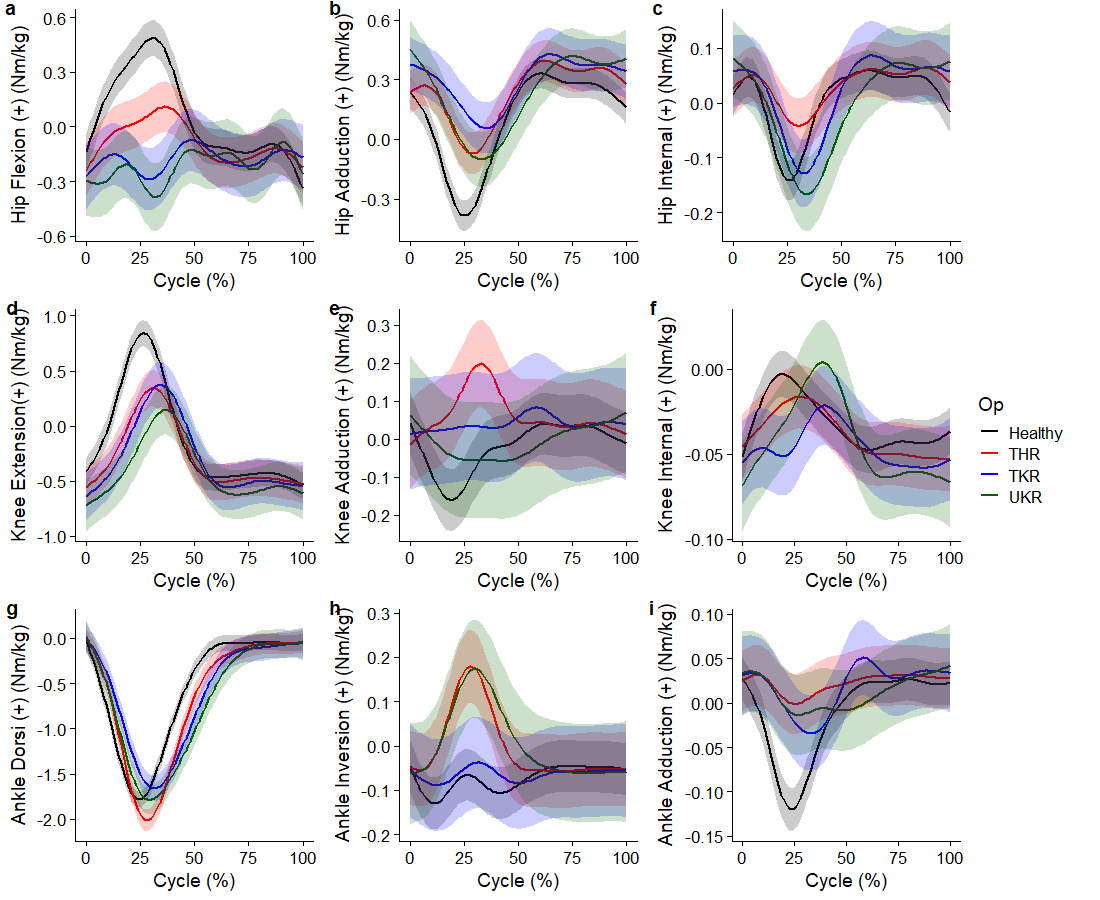

#### Figure SM15. Bilateral hopping – cycle reflects the stride phase

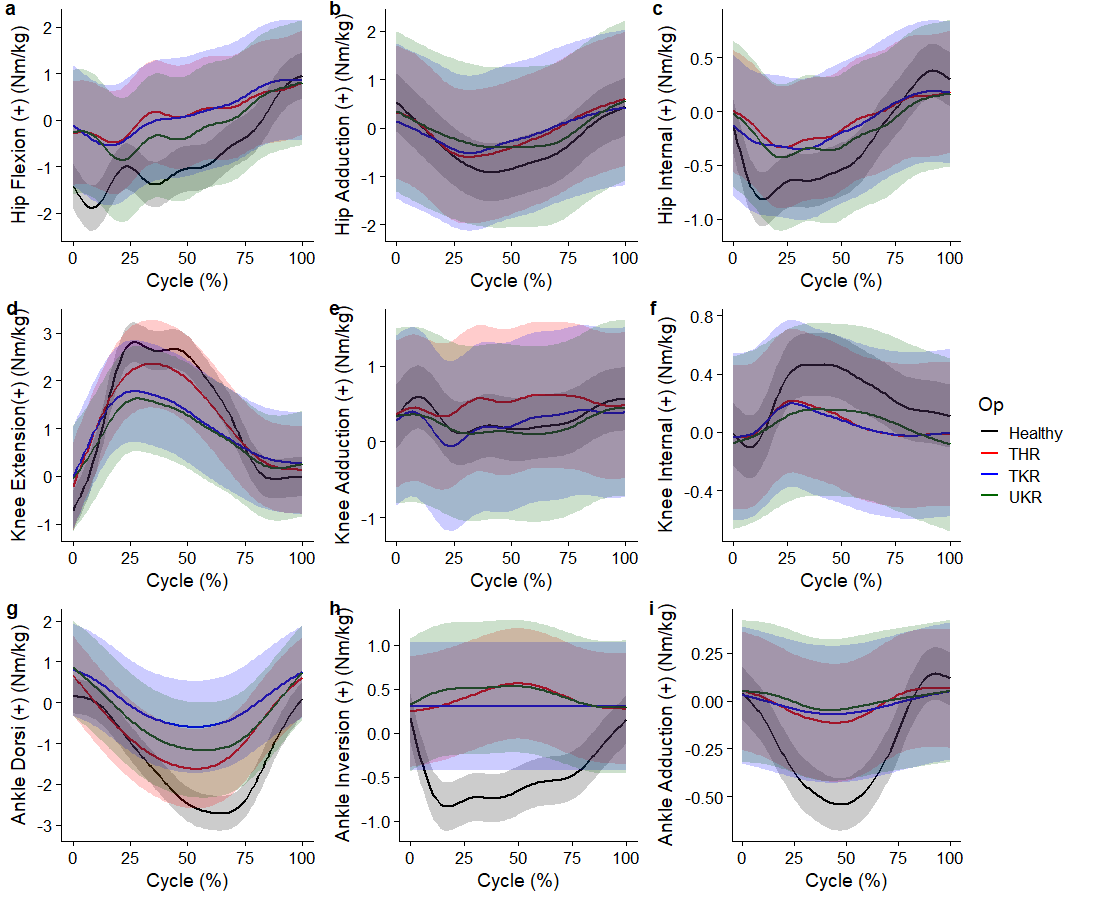

#### Figure SM16. Change of direction 45° – cycle reflects the stance phase

### Joint moment difference plots

Figures below reflect the time-varying waveforms of the pairwise difference between groups for each point on the movement cycle during various activities. Lines reflect the point estimate difference, whilst error clouds reflect 95% confidence intervals. Horizontal lines above waveforms reflect periods of statistically significant difference (i.e. non-zero cross of 95% confidence intervals).

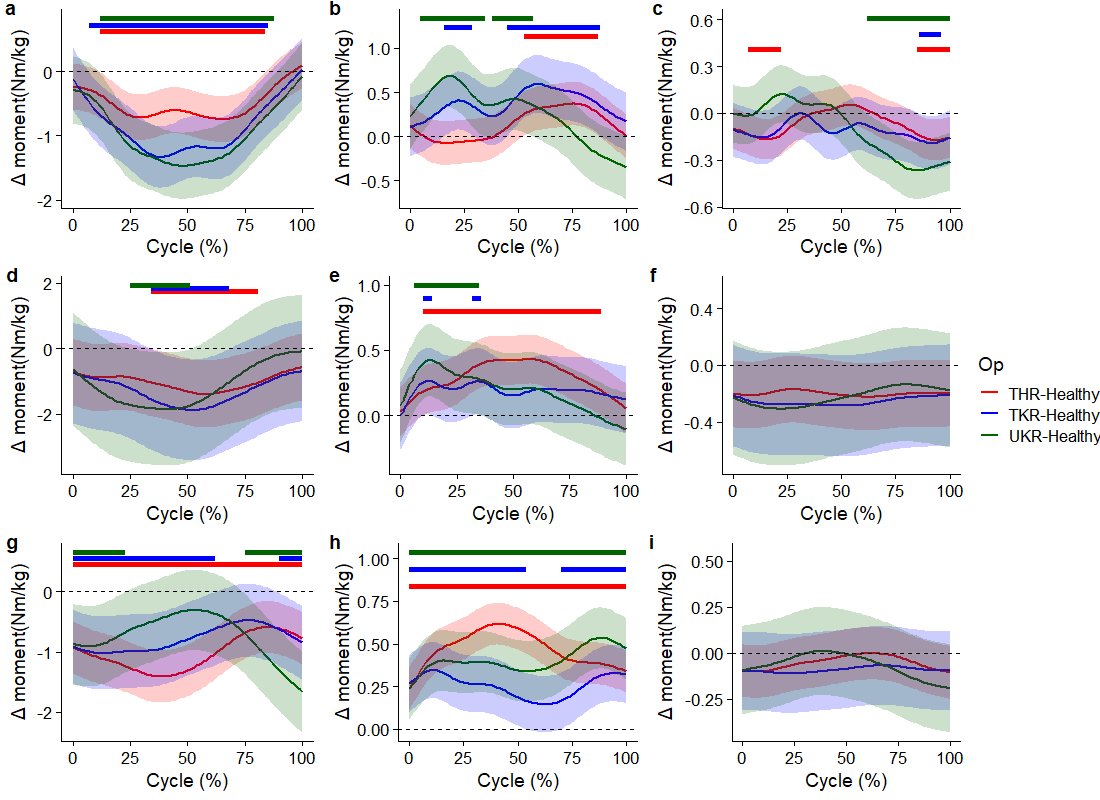

#### Figure SM17. Running – cycle reflects the stance phase.

(a) Hip flexion (+ = greater flexion in target), (b) Hip adduction (+ = greater adduction in target), (c) Hip internal rotation (+ = greater internal rotation in target), (d) Knee flexion (+ = greater extension in target), (e) Knee adduction (+ = greater adduction in target), (f) Knee internal rotation (+ = greater internal rotation in target), (g) Ankle Dorsi (+ = greater dorsiflexion in target), (h) Ankle inversion (+ = greater inversion in target), (i) Ankle Adduction (+ = greater adduction in target).

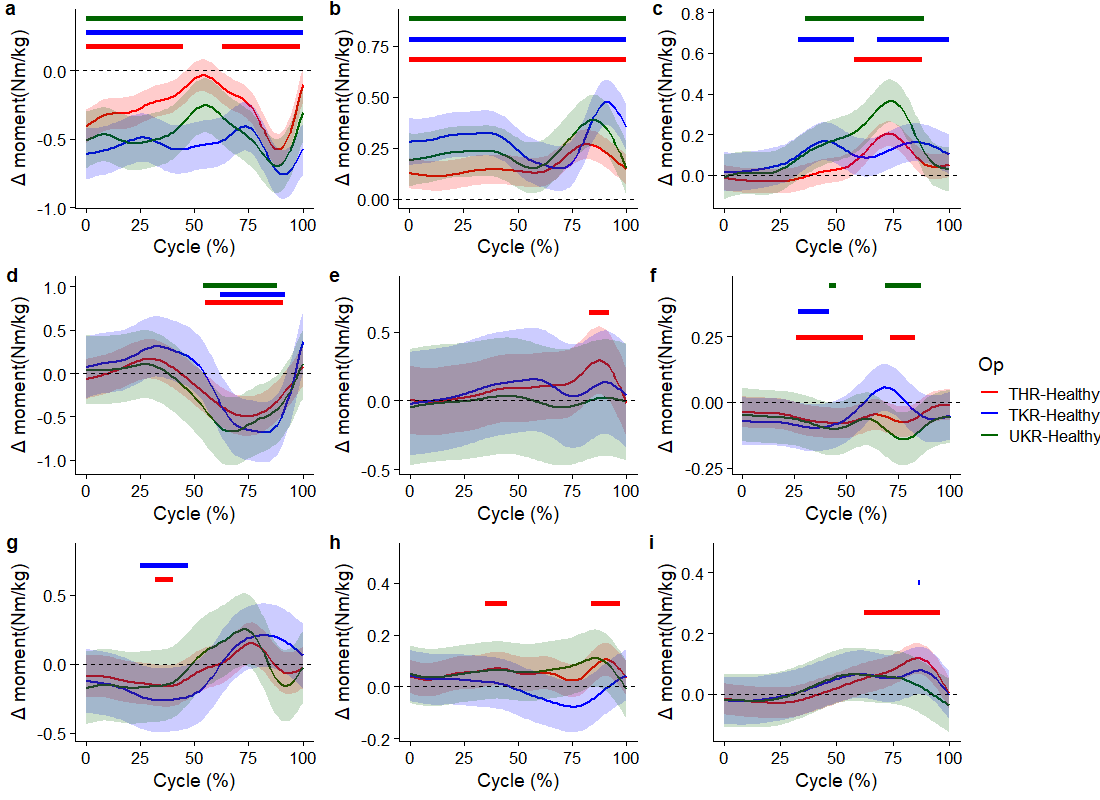

#### Figure SM18. Countermovement jumps – cycle reflects from the lowering of the centre of mass to toe-off

(a) Hip flexion (+ = greater flexion in target), (b) Hip adduction (+ = greater adduction in target), (c) Hip internal rotation (+ = greater internal rotation in target), (d) Knee flexion (+ = greater extension in target), (e) Knee adduction (+ = greater adduction in target), (f) Knee internal rotation (+ = greater internal rotation in target), (g) Ankle Dorsi (+ = greater dorsiflexion in target), (h) Ankle inversion (+ = greater inversion in target), (i) Ankle Adduction (+ = greater adduction in target).

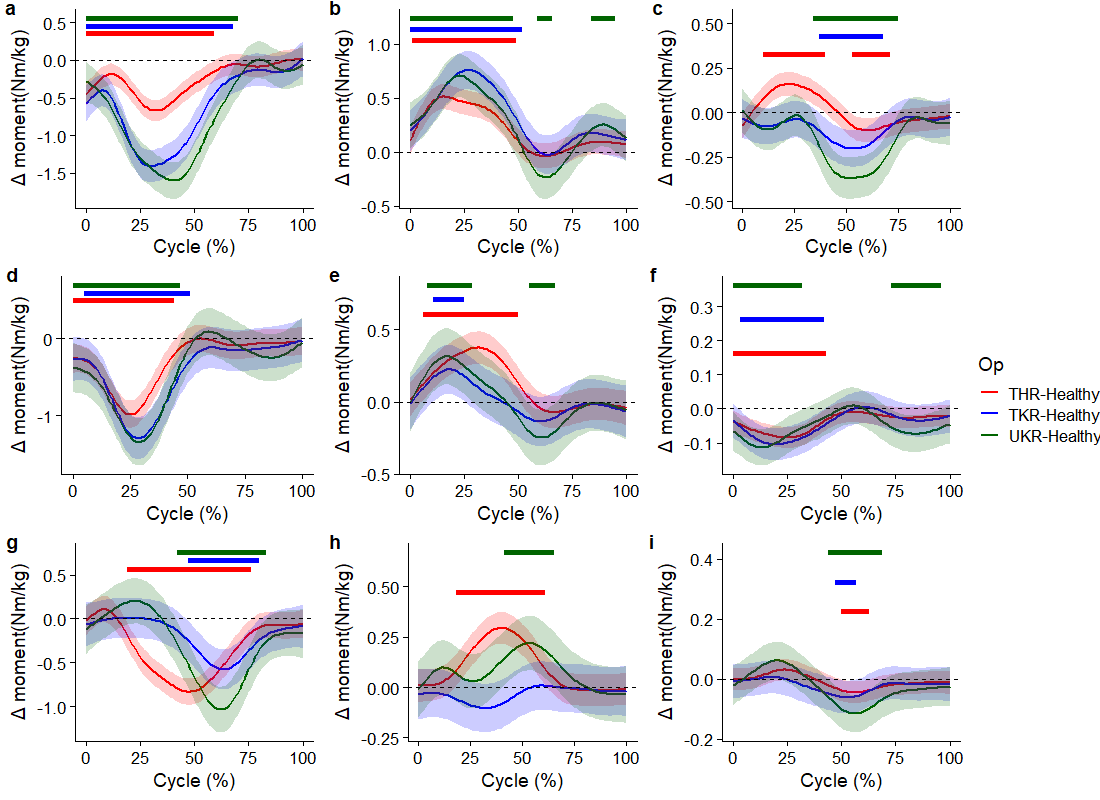

#### Figure SM19. Unilateral hopping – cycle reflects the stride phase

(a) Hip flexion (+ = greater flexion in target), (b) Hip adduction (+ = greater adduction in target), (c) Hip internal rotation (+ = greater internal rotation in target), (d) Knee flexion (+ = greater extension in target), (e) Knee adduction (+ = greater adduction in target), (f) Knee internal rotation (+ = greater internal rotation in target), (g) Ankle Dorsi (+ = greater dorsiflexion in target), (h) Ankle inversion (+ = greater inversion in target), (i) Ankle Adduction (+ = greater adduction in target).

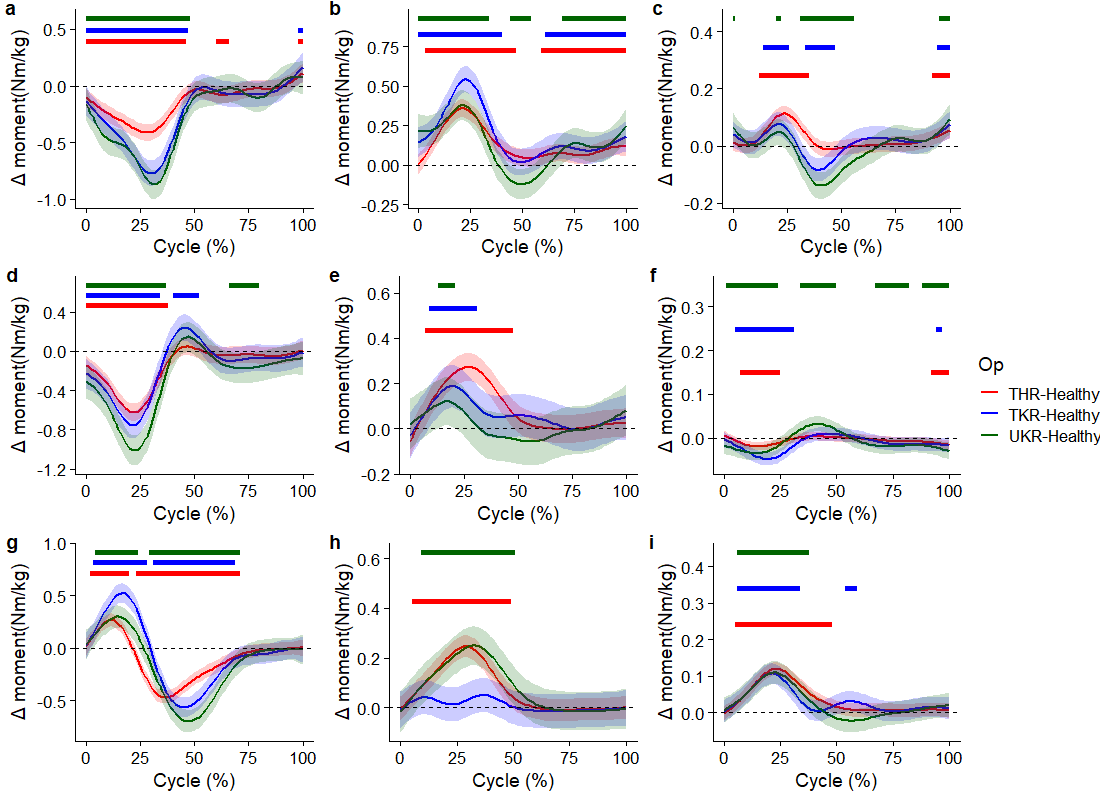

#### Figure SM20. Bilateral hopping – cycle reflects the stride phase

(a) Hip flexion (+ = greater flexion in target), (b) Hip adduction (+ = greater adduction in target), (c) Hip internal rotation (+ = greater internal rotation in target), (d) Knee flexion (+ = greater extension in target), (e) Knee adduction (+ = greater adduction in target), (f) Knee internal rotation (+ = greater internal rotation in target), (g) Ankle Dorsi (+ = greater dorsiflexion in target), (h) Ankle inversion (+ = greater inversion in target), (i) Ankle Adduction (+ = greater adduction in target).

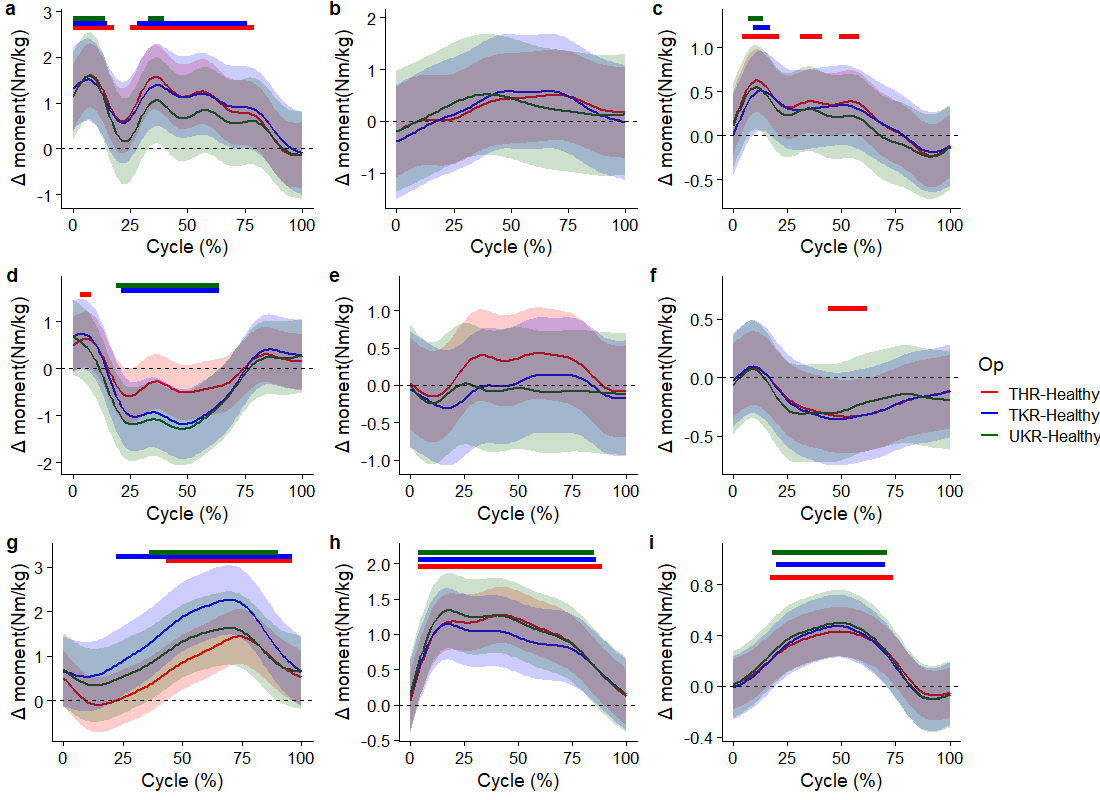

#### Figure SM21. Change of direction 45° – cycle reflects the stance phase

(a) Hip flexion (+ = greater flexion in target), (b) Hip adduction (+ = greater adduction in target), (c) Hip internal rotation (+ = greater internal rotation in target), (d) Knee flexion (+ = greater extension in target), (e) Knee adduction (+ = greater adduction in target), (f) Knee internal rotation (+ = greater internal rotation in target), (g) Ankle Dorsi (+ = greater dorsiflexion in target), (h) Ankle inversion (+ = greater inversion in target), (i) Ankle Adduction (+ = greater adduction in target).
